# Optimality of Maximal-Effort Vaccination

**DOI:** 10.1101/2022.05.12.22275015

**Authors:** Matthew J. Penn, Christl A. Donnelly

**Affiliations:** Department of Statistics, University of Oxford, St Giles’, Oxford, OX1 3LB, United Kingdon; Department of Infectious Disease Epidemiology, Imperial College London, South Kensington Campus, London, SW7 2AZ, United Kingdom; Pandemic Sciences Institute, University of Oxford, Roosevelt Drive, Oxford, OX3 7DQ, United Kingdon

**Keywords:** Vaccination, Epidemiology, Epidemics, SIR Modelling

## Abstract

It is widely acknowledged that vaccinating at maximal effort in the face of an ongoing epidemic is the best strategy to minimise infections and deaths from the disease. Despite this, no one has proved that this is guaranteed to be true if the disease follows multi-group SIR (Susceptible-Infected-Recovered) dynamics. This paper provides a novel proof of this principle for the existing SIR framework, showing that the total number of deaths or infections from an epidemic is decreasing in vaccination effort. Furthermore, it presents a novel model for vaccination which assumes that vaccines assigned to a subgroup are distributed randomly to the unvaccinated population of that subgroup. It suggests, using COVID-19 data, that this more accurately captures vaccination dynamics than the model commonly found in the literature. However, as the novel model provides a strictly larger set of possible vaccination policies, the results presented in this paper hold for both models.

## 1 Introduction

The COVID-19 pandemic has illustrated the importance of quickly implementing vaccination policies which target particular groups within a population Fitzpatrick and Galvani (2021). The difference in final infections between targeted policies and uniform distribution to the entire population can be significant Castro and Singer (2021), Estadilla et al (2021) and so it is important that the models underlying these decisions provide realistic predictions of the outcomes of different policies.

One of the most commonly used models to forecast epidemics is the multi-group SIR (Susceptible-Infected-Recovered) model Ram and Schaposnik (2021), Acemoglu et al (2021), Kuniya (2019). This model divides the population into different groups based on characteristics such as age or occupation. Each group is then further sub-divided into categories of susceptible, infected and recovered. Where vaccination does not give perfect immunity, further sub-categorization based on vaccination status can also be used Kuga and Tanimoto (2018), as will be done in this paper.

While many other approaches have been developed either by adding compartments to the SIR framework Moore et al (2021) or using completely different models such as networks Chen and Sun (2014) or stochastic simulations Ball and Lyne (2002), the multi-group SIR model remains popular because of its comparatively small number of parameters and its relatively simple construction and solution. In this paper, attention will thus be restricted to the multi-group SIR model, although it would be beneficial for future work to consider a wider range of disease models.

There are two general frameworks that are used to model optimal vaccination policies in a resource-limited setting. The first, used in papers such as Hill and Longini Jr (2003) and Becker and Starczak (1997), seeks to reduce the reproduction number, *R*_0_ of the epidemic as much as possible by vaccinating before infections arrive in a population. It is simple to show that in this case, one should use all of the vaccinations available, and so this problem will not be considered further in this paper.

The second framework, used in papers such as Acemoglu et al (2021) and Hansen and Day (2011) aims to minimise the total cost of an epidemic. This is the framework that will be discussed in this paper. The “cost” of an epidemic is, in general, defined to be the number of deaths (or equivalently, infections), with many papers also considering the cost of vaccination alongside the cost of other control measures, such as isolation, lockdown or treatment Fu et al (2022).

One important principle which underlies all of these vaccination policies is the acceptance that giving people their first dose of vaccine as soon as possible reduces the number of infections. Of course, this only holds when the timescale considered is sufficiently short for effects such as waning immunity and disease seasonality to be negligible, and a more complicated framework would be needed to model these effects. However, the acceptance of at least short-term optimality of maximal vaccination effort has been highlighted in the COVID-19 pandemic response, as countries began their vaccination programs as soon as vaccines became available Mathieu et al (2021).

To the best of the authors’ knowledge, no one has provided a mathematical proof that in a general, multi-group SIR model with imperfect vaccination, it is always best to vaccinate people as early as possible. Of course, it is not difficult to create a conceptually sound justification - vaccinating more people means that fewer people will catch the disease which will reduce the total number of infections. However, the SIR model is an approximation of the process of a disease spreading, and so it is important that it obeys this principle for all physical parameter values and vaccination policies.

Some special cases of the theorem presented in this paper have been previously proved in the literature. In particular, a significant number of papers have considered the optimal vaccination policy for a homogeneous population, with Abakuks (1972) first proving that, in this case, it is optimal to vaccinate at maximal effort (if one ignores the cost of vaccination). This proof held for vaccination policies that were finite sums of point mass “impulse” vaccinations, and has been generalised by papers such as Hansen and Day (2011), Zaman et al (2008), Morton and Wickwire (1974) and Zhou et al (2014) to a much wider class of vaccination policies, although the proof was still restricted to a single group and to perfect vaccination. Moreover, Hansen and Day (2011) notes that the case of imperfect vaccination (where vaccinated individuals can still get infected, although at a lower rate) remained a topic of open investigation, and so it can not easily be solved using the same methods presented in these papers. A slight extension is made in Duijzer et al (2018) where it is shown that maximal effort is optimal in the case of perfect vaccination of any number of disconnected groups, but the full problem is still far from understood.

The general method of proof in the literature relies on Pontryagin’s Maximum Principle, which is difficult to apply to multi-group models due to the more complex structure of the equations. It is simple to characterise the solution in terms of the adjoint variables, as is done in Zhang et al (2020) and Zavrakli et al (2021) for a two-group model with imperfect vaccination, in Boutayeb et al (2021) for a general *n*-group model with perfect vaccination and in Lee et al (2012) for a six-group model with imperfect vaccination. However, determining whether this solution corresponds to the maximal effort solution in the case of zero vaccination cost requires the analysis of the adjoint ODE system, which is often just as complicated as the original disease model. In particular, the fact that vaccinated people need to be no more infectious, no more susceptible and be infected for no more time than unvaccinated people means that any analysis of the adjoint system would be complicated, as the properties of all the constituent parameters would need to be used.

Thus, in this paper, a novel approach is developed. Rather than attempting to use the general optimal control theory methodology, the specific structure of the SIR equation system is exploited. Using this, an inequality is derived which shows that if a given vaccination policy, ***Ũ*** vaccinates each individual at least as early as another vaccination policy, ***U***, then the latter policy will lead to at least as many deaths (or equivalently, infections) as the former. As well as providing a constraint on the optimal solution, this theorem also highlights important structural properties of the model, as it shows that the number of deaths is everywhere non-increasing in the vaccination rates, rather than this just holding near the optimal solutions.

Also introduced in this paper is a more general, and seemingly more accurate, model of vaccination than the one normally used in the literature. The one that is typically used (in almost all papers cited in this work such as Hansen and Day (2011), Zaman et al (2008) and Kar and Batabyal (2011)) assumes that the entire population is randomly vaccinated at a controllable rate. In practice, this is unrealistic, as once a person has been vaccinated once, that information is available to policy-makers so they can ensure that only the unvaccinated are given new doses (assuming that only one dose of the vaccine is required). There can be significant differences between the optimal vaccination policy according to the two models, and so it seems that the model introduced in this paper should be strongly considered for new optimal vaccination studies.

However, it is shown in this paper that, in the context of the theorems proved, modifying the model so that vaccines are assigned randomly to the unvaccinated population provides a strictly larger set of possible vaccination policies. In particular, this means that the results proved here also apply to the more commonly-used case, and so can also be used by those following the more commonly-used vaccination model.

Alongside proving that the final infected, recovered and dead populations are non-increasing with increased and earlier vaccination effort, some cautionary contradictions to perhaps intuitive conjectures are also provided which show the importance of mathematical proof instead of simply intuition. In particular, it is shown that increased vaccination (under this model) can lead to, at a fixed finite time of the simulation, higher infection rates or a higher death count, despite the longer-term better performance of this policy. Indeed, it is results similar to these which make the proof of the optimality of maximal effort difficult, as it means that one must be very careful when constructing the inequalities that do hold for all models.

## 2 Modelling

### 2.1 Disease Transmission and Vaccination Model

Suppose that the population is divided into *n* subgroups, such that population of people in group *i* is *N*_*i*_ and define

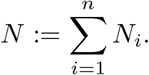

Define the compartments of people as follows, for *i* = 1, …, *n*:

*S*_*i*_ :=Number of people that are in group *i*, are susceptible, and are unvaccinated,

*I*_*i*_ :=Number of people that are in group *i*, are currently infected, and were infected while unvaccinated,

*R*_*i*_ :=Number of people that are in group *i*, are recovered or dead, and were infected while unvaccinated,

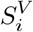:=Number of people that are in group *i*, are susceptible and are vaccinated,

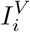:=Number of people that are in group *i*, are infected and were infected after being vaccinated,

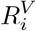:=Number of people that are in group *i*, are recovered or dead and were infected after being vaccinated.

It is assumed that there is a record of people who have received a vaccination and that protection from vaccination does not decay over time, so that no one is vaccinated more than once. Thus, if a total number, *U*_*i*_(*t*)*dt*, of people in group *i* are given vaccines in a small time interval (*t, t* + *dt*), and these vaccines are distributed randomly to the unvaccinated population in group *i*, the total population of susceptibles given vaccines in group *i* is

*U*_*i*_(*t*)*dt*×P(A person in group *i* is in *S*_*i*_ | A person is in group *i* is unvaccinated) which is equal to

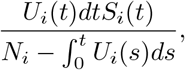

as 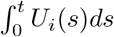 is the total population that are in group *i* and have been vaccinated before time *t*. For the remainder of this section, this vaccination model will be referred to as the “unvaccinated-only” model

This results in the following model, based on SIR principles

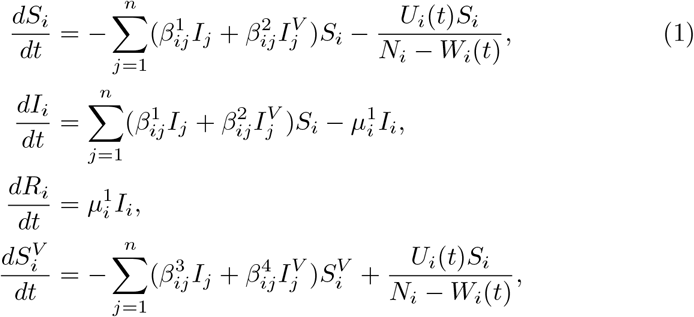

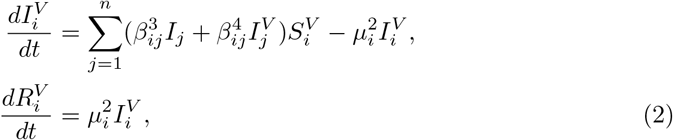

where

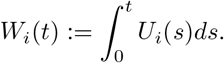

Here, 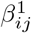 represents transmission from the unvaccinated members of group *j* to the unvaccinated members of group *i*, 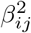 represents transmission from vaccinated members to unvaccinated members, 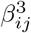 represents transmission from vaccinated members to unvaccinated members and 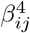 represents transmission from vaccinated members to vaccinated members. Additionally, 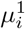 represents the infectious period of unvaccinated infected members in group *i* while 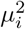 represents the infectious period of vaccinated members. Note that the superscript denotes different parameter values, so that 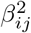 is not necessarily the square of 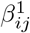.

To ensure that vaccination is “locally effective” (that is, a vaccinated individual is no more likely to transmit or be infected by the disease, and is infectious for no longer than an unvaccinated individual in the same sub-group), and that the parameters are epidemiologically feasible, the following constraints are imposed:

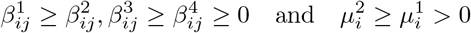

Note that there is no constraint on the ordering of 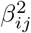 and 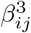. It is assumed for convenience that all variables except the *S*_*i*_ and *I*_*i*_ are initially zero and that all initial conditions are non-negative.

The equations (1) - (2) sum to zero on the right-hand side, and so for each *i*,

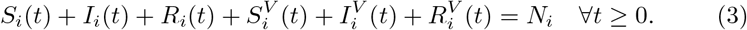

It will be assumed that the populations and parameters have been scaled such that *N* = 1, Finally, it is assumed that

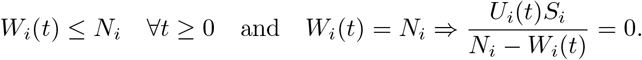

to ensure feasibility of the vaccination policies.

### 2.2 Fully Random Vaccination

A more common model of vaccination in the literature is the “fully random” vaccination model (Hansen and Day (2011), Zaman et al (2008) and Kar and Batabyal (2011)), where vaccinations assigned to each subgroup are given randomly to all members of that subgroup. That is, the equation (1) becomes

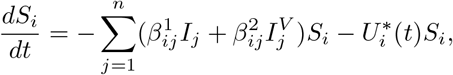

where 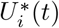 is the vaccination rate in this model.

Dimensional analysis reveals that that 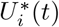 has dimension 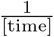, unlike *U*_*i*_(*t*) (which has dimension 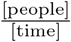). Thus, *U*_*i*_(*t*)*dt* is the number of vaccines given in a small time interval [*t, t* + *dt*], while 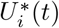 instead is the proportion of the population that are given vaccines in this interval. Thus, the number of vaccinations assigned by the fully random model in the small time interval [*t, t* + *dt*] is 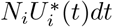. This model then corresponds to random vaccination as

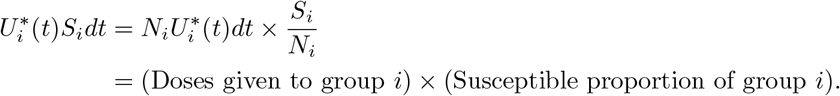

As in Zaman et al (2008), 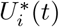 can equivalently be interpreted as the percentage of susceptibles being vaccinated per unit time. However, even with this interpretation, Zaman et al (2008) constrains 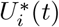 to be less than a fixed, constant value. This is still in line with the random vaccination interpretation, as this constraint could again be interpreted as simply the number of randomly-assigned vaccines that are “successful” in being given to susceptibles, given an overall constant rate of vaccination to the whole population.

### 2.3 Justification of the Unvaccinated-Only Model

A significant problem with the fully random model is that it assumes that the number of vaccines wasted when vaccinating a population grows very quickly as more people are vaccinated. For example, in a (mostly susceptible) population that has 50% vaccination coverage, then it assumed that it will take twice as much effort to vaccinate the same number of people as at the start of program. This seems unrealistic given the vaccination uptake for COVID-19, where demand remained high until a large proportion of the population had been vaccinated, as shown in Ritchie et al (2020). Moreover, Aubrey et al (2022) shows that in Canada, the number of wasted vaccines was 2.6% of the total given out when their adult population had been mostly vaccinated (at least once) in November 2021, and so it is not the case that the vaccine wastage scales in the way predicted by the fully random model.

Figure 1, made from the data used in Ritchie et al (2020) and created in Mathieu et al (2021) gives an explicit example of this. It compares the median time taken (in the 27 countries from the dataset that recorded their data in the same way) to give a first vaccine dose to different 10% percentiles of the population that were over 80 years old in each country. As a baseline, it uses the time taken to move from 20% vaccination coverage of this age group to 30% coverage (to avoid errors from the very small but rapidly growing initial vaccine supply). The median values shown in the figure are the median of the times

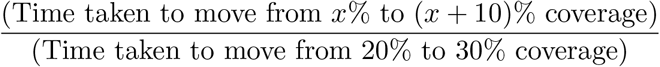

across the 27 different countries, where *x* denotes the value on the *x* axis. Assuming that vaccination effort was roughly constant in the period during which these vaccinations occurred (which is not a completely accurate assumption, but in general, those aged over 80 years were given their first doses in a short period of time), one would then expect that these proportions would be equal to 1 for all *x* under the unvaccinated-only model. However, under the fully random model, significantly more effort is required to vaccinate people once the total percentage vaccinated is higher. Indeed, if *X* is the unvaccinated percentage then one has (assuming 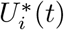 is a constant)

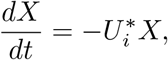

and hence

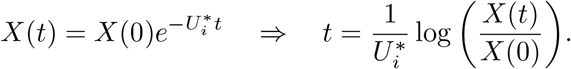

**Fig. 1.**
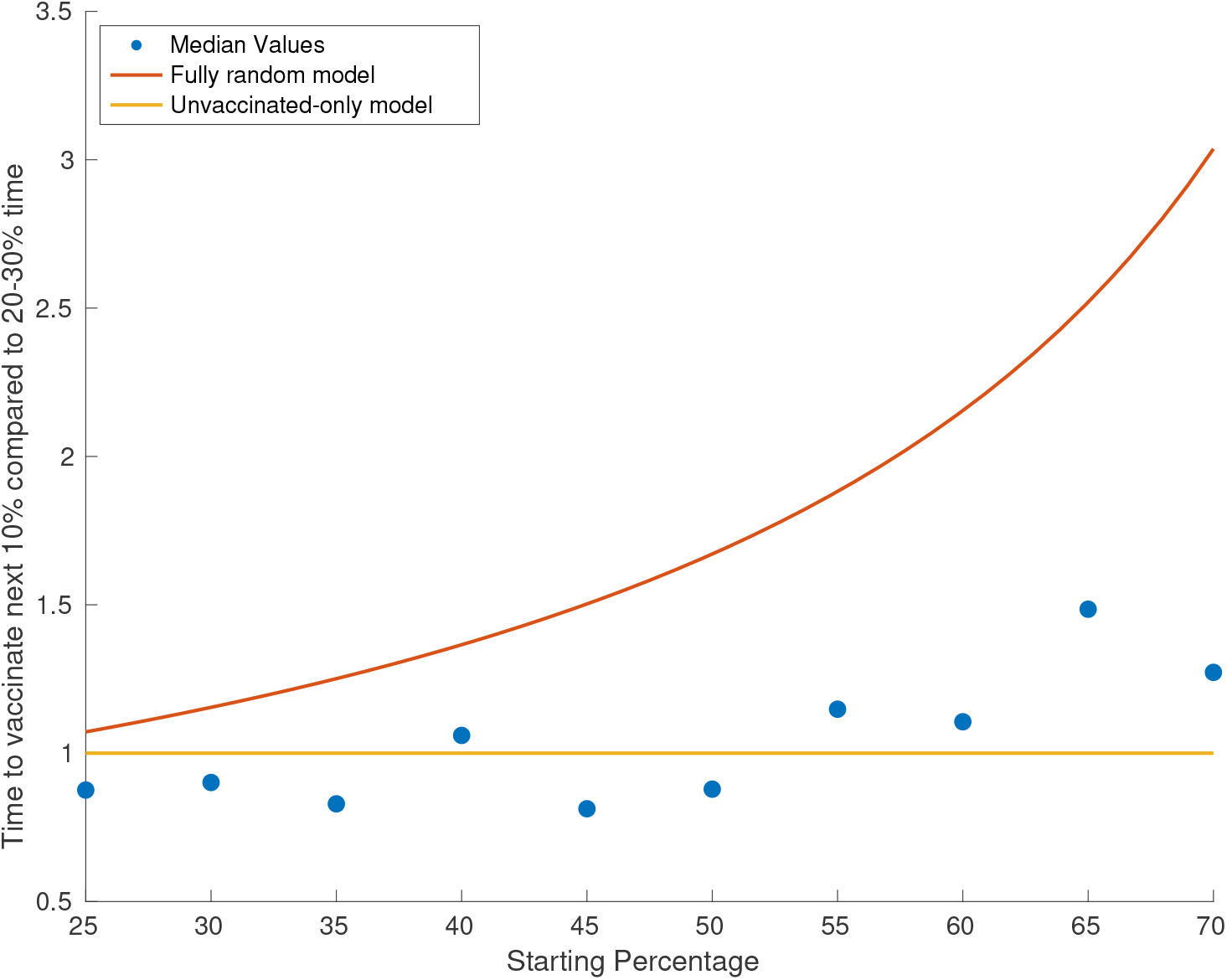
A comparison of the expected time to vaccinate the next 10% of those aged over 80 years according to the fully random model, unvaccinated-only model and median values from data taken from 27 countries.

These predicted times according to the random model are given by the red line in Figure 1.

It is clear from Figure 1 that the constant-time-interval model provides a much better prediction of the median time intervals from the data. Of course, it is important to model the fact that a significant proportion of each group in the population will remain unvaccinated, and that rates of uptake will decrease as the number of unvaccinated people decreases, evidence of which is apparent in Figure 1. However, both models could easily be modified to (more accurately) incorporate the fact that not everyone will get vaccinated by constraining *W*_*i*_(*t*) ≤ *α*_*i*_*N*_*i*_ for some *α*_*i*_ *<* 1. Furthermore, the demand could be modelled by introducing some general constraint *U*_*i*_(*t*) ≤ *K*(*W*_*i*_(*t*), *t*) which would again more accurately model this phenomenon.

Thus, it seems that the unvaccinated-only model does give a more accurate representation of a vaccination program than the more traditional random model, although more detailed analysis would be needed to justify this further. Therefore, future studies should consider carefully their choice of vaccination function to ensure that their results are as practically applicable as possible.

### 2.4 The Importance of Choosing the Correct Model

The distinction between the two models is important, as there are many examples where the two models will give significantly different solutions to the optimal vaccination problem. Consider a two-group case with perfect vaccination (so 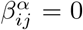 for *α >* 1) with *N*_1_ = *N*_2_ = 1 (which means that 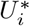 and *U*_*i*_ are constrained in the same way). Then, suppose the parameters are

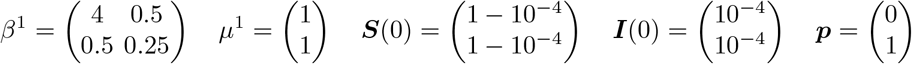

where ***p*** is the case fatality ratio in each group. Suppose that the objective is to minimise the final number of deaths, given by *R*_2_(∞). Moreover, suppose that the overall vaccination rate is given by

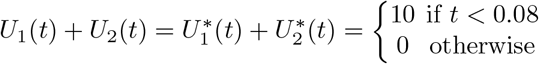

so that vaccination takes place almost instantly. Suppose that one has the choice between vaccinating exactly group 1 or group 2 at maximum rate until the supply runs out. Then, under the random vaccination model, it is not possible to reach herd immunity by vaccinating the infectious group 1, and so the optimal strategy is to vaccinate group 2. However, in the unvaccinated-only model it *is* possible to reach herd immunity and the optimal strategy is to vaccinate group 1. Indeed, if the random model is used to make the decision, but the unvaccinated-only model is the actual population response to vaccination, then the wrong decision is made and the number of deaths increases from 7 × 10^−4^ to 0.08. Thus, it is clearly important to ensure that the correct model is chosen.

### 2.5 Globality of Results in This Paper

However, the principle of maximal vaccination effort will hold for both models of vaccination, which can be shown as follows. The two models are equivalent whenever

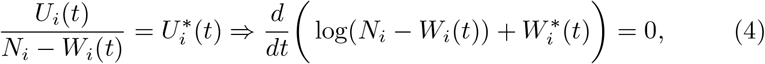

where

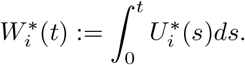

Thus, by integrating (4), and noting that 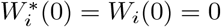

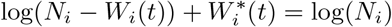

and so

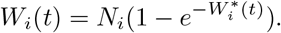

Thus, given any fully random vaccination policy ***U*** ^*^, it can be replaced by a unvaccinated-only policy ***U*** (although the converse does not hold as *W*_*i*_(*t*) = *N*_*i*_ requires 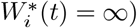).

Moreover, note that 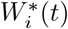 is increasing in *W*_*i*_(*t*). Thus, if a pair of fully random vaccination policies ***U*** and ***Ũ*** satisfy 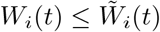 then this inequality is preserved by the corresponding unvaccinated-only policies as 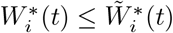. This property means that the theorems proved in this paper will hold for both models (as they will be proved using an “unvaccinated only” model).

## 3 Optimisation Problem

Now that the model has been formulated, it is possible to set up the optimisation problem that will be considered in the remainder of this paper.

### 3.1 Constraints on *U*_*i*_(*t*)

In order to assist the proof of the theorems, it is necessary to make some (unrestrictive) assumptions on the vaccination rates, *U*_*i*_(*t*).

Firstly, there are the physical constraints that for each *i* ∈ *{*1, …, *n}*

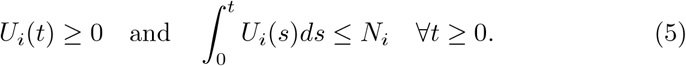

It is also necessary that *U*_*i*_(*t*) is within the class of functions such that solutions to the model equations exist and are unique. Discussion of the exact conditions necessary for this to hold is outside the scope of this paper. However, from the Picard-Lindelöf Theorem Collins (2006), a sufficient condition for this is that *U*_*i*_(*t*) is a piecewise Lipschitz continuous function. While this is not a necessary condition, this illustrates that this assumption will hold for a large class of functions. However, it will be helpful throughout the course of the proof to explicitly assume two conditions on *U*_*i*_(*t*) - namely, that it is bounded and that it is Lebesgue integrable on ℜ for each *i*.

For the remainder of this paper, define the set of feasible vaccination policies, *C*, is the set of functions ***U*** satisfying (5) such that unique solutions to the model equations exist with these functions as the vaccination policy and such that each *U*_*i*_(*t*) is bounded and Lebesgue integrable on ℜ.

### 3.2 Optimisation Problem

The aim is to choose the vaccination policy ***U*** ∈ *C* such that the total number of deaths (or any linear function of the infections in each subgroup) is minimised while meeting additional constraints on vaccine supply and vaccination rate. It is assumed that the maximal rate of vaccination at time *t* is *A*(*t*) and that there is a total (non-decreasing) supply of *B*(*t*) vaccinations that has arrived by time *t*. Thus, for each *i, U*_*i*_(*t*) is constrained to satisfy

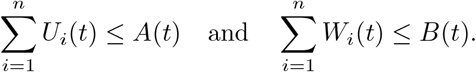

It is assumed that each infection of unvaccinated people in group *i* is weighted by some *p*_*i*_ (this could be interpreted as a probability of death, hospitalisation or, if *p*_*i*_ = 1, simply counting the number of infections). Moreover, it is assumed that the infection is no more serious for those that have been vaccinated, so that the weighting of an infection of a vaccinated person in group *i* is *p*_*i*_*κ*_*i*_, where *κ*_*i*_ ≤ 1. Thus, the objective function is

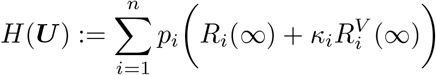

where, for example

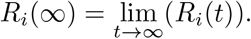

Note these limits exist as *R*_*i*_ is non-decreasing and bounded by Lemma B.3. Hence, the optimisation problem is

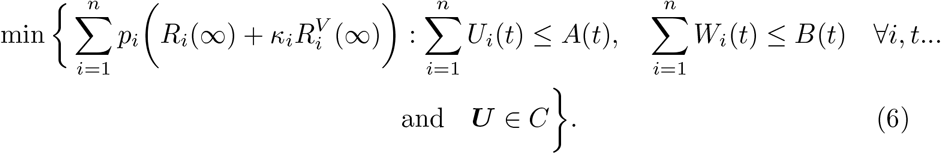

## 4 Main Results

The main results of this paper are as follows. Firstly, it is shown that the objective function is non-increasing in vaccination effort.

### Theorem 1.

*Suppose that* ***U***, ***Ũ*** ∈ *C. Suppose further that for each i* ∈ *{*1, …, *n} and t* ≥ 0

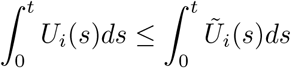

*Then*

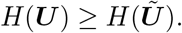

Then, it is shown that if an optimal solution exists, there is an optimal maximal effort solution.

### Theorem 2.

*Suppose that B is differentiable, and that there is an optimal solution* ***U*** *to (6). Then, define the function*

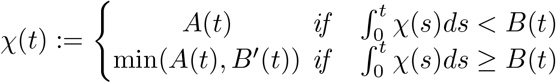

*and suppose that χ*(*t*) *exists and is bounded. Then, there exists an optimal solution* ***Ũ*** *to the problem (6) such that*

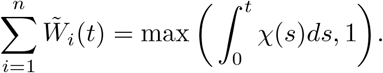

*Moreover, if χ*(*t*) *is continuous almost everywhere, there exists an optimal solution* ***Ũ*** *such that*

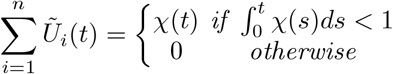

It is perhaps concerning to the reader that the existence of *χ* is left as an assumption in this theorem. However, while the exact conditions on its existence are beyond the scope of this paper, it certainly exists for a wide class of functions *A*(*t*) and *B*(*t*), as proved in Lemma A.11.

Finally, it is shown that this principle still holds if the cost of vaccination is considered.

### Theorem 3.

*Under the assumptions of Theorem 2, consider a modified objective function* ℋ *given by*

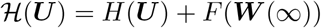

*for any function F. Then, with χ defined to be the maximal vaccination effort as in Theorem 2, there exists an optimal solution* ***Ũ*** *such that, for some τ* ≥ 0

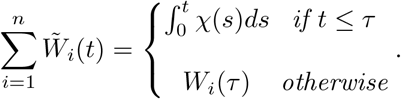

*Moreover, if χ is continuous almost everywhere, then there is an optimal solution* ***Ũ*** *such that*

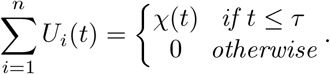

## 5 Proof

Before beginning the main proof, it is helpful to note some fundamental results about the SIR equations that will be used throughout. Namely, for each *i* ∈ *{*1, …, *n}* and *t* ≥ 0

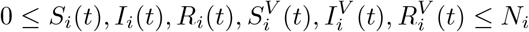

and

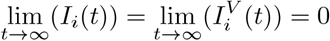

These results are proved in Lemmas B.3 and B.4.

It is first useful to define

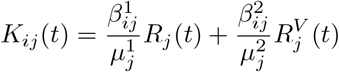

and

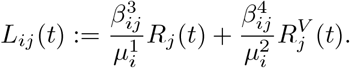

Then, the following propositions hold.

### 5.1 An Inequality for *K*_*ij*_ and *L*_*ij*_

Note that the proof of this proposition requires a significant amount of algebra, and the majority of it has hence been left to lemmas which can be found in Appendix A. However, the key logic of the proof will be presented here.

Also, note that in this paper, a step function is defined to be a function that is piecewise constant on any *bounded* interval of ℜ. Thus, it may have infinitely many discontinuities, but only finitely many in any bounded interval. This differs from the definition used in some other papers (which impose that a step function is piecewise constant on ℜ).

#### Proposition 1.

*Suppose that U*_*i*_(*t*) *and Ũ*_*i*_(*t*) *are right-continuous step functions. Moreover, suppose that*

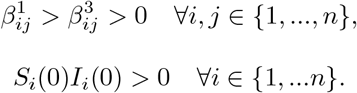

*and that*

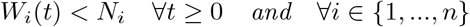

*Then*,

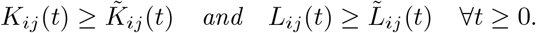

**Proof:** Suppose that the proposition does not hold. Hence, one can define

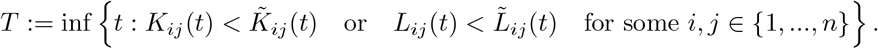

Then, there exists some *b* ∈ {1, .., *n*} and some real constants *κ* and *η* such that the following system of inequalities holds at time *T* :

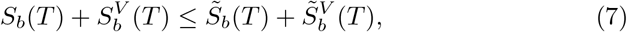

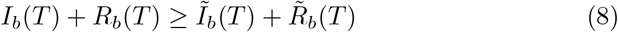

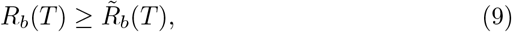

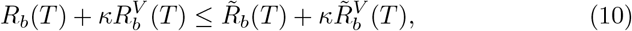

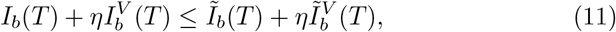

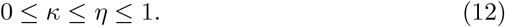

The derivations of inequalities (7) - (12) are found in Lemmas A.2 - A.5. Moreover,

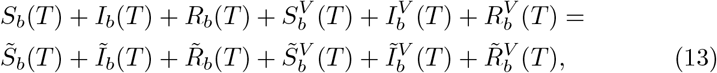

which comes from (3). Note that (10) in fact holds to equality in this case, but this is not necessary for the proof (and later, the same system will be considered where such an equality is not guaranteed).

By Lemma A.6, the system (7) - (13) implies that

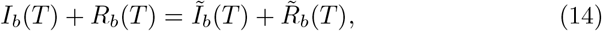

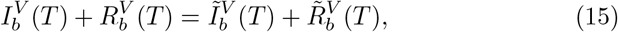

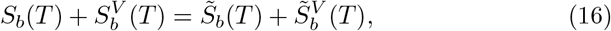

If *T >* 0, then Lemma A.7 can be used to show that

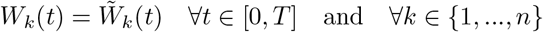

while if *T* = 0 then this is immediate. Thus, the two ODE systems are the same up to time *T*, which means that all variables (in all groups) are equal at time *T*.

From this point, the proof of Proposition 1 can be completed by considering the behaviour of the system at time *T* + *δ* for small *δ*. For sufficiently small *δ, U*_*i*_(*t*) and *Ũ*_*i*_(*t*) are constant on [*T, T* + *δ*] (as they are step functions) and this condition on *δ* will be assumed for the remainder of this proof

Define functions 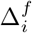 to be

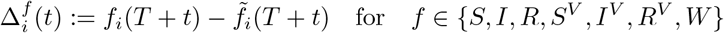

and note that

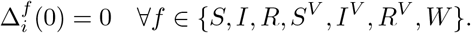

Then, by Lemma A.8, for *t* ∈ [0, *δ*] and any real numbers *x* and *y*

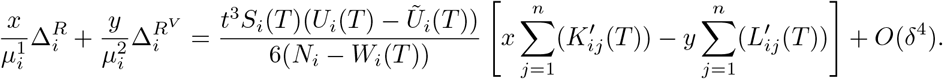

Hence, by Lemma A.9,

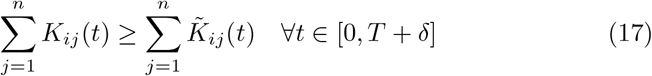

and

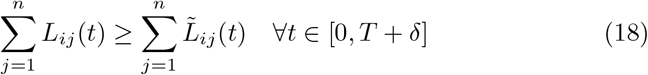

for sufficiently small *δ*.

Now, by the definition of *T*, there exists some *t* in [*T, T* + *δ*] such that, for some *a, b*

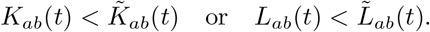

Indeed, from Lemma A.10, there exists some *t* ∈ (*T, T* + *δ*) such that

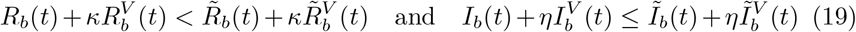

for some

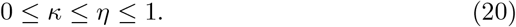

Now, by Lemmas A.2 - A.4 (which only require the properties (17) and (18)), the system of inequalities (7)-(9) holds for group *b* at time *t*. These can be combined with (19), (20) and (13) to use Lemma A.6, showing

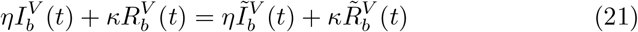

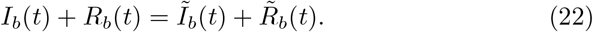

By adding the inequalities in (19) together,

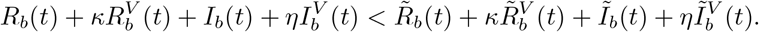

Then, (21) and (22) show that this must in fact be an equality which is a contradiction. Thus, *t* cannot exist. This provides a contradiction to the definition of *T*, and hence finishes the proof of Proposition 1.

It is now possible to prove Theorem 1 under the extra restrictions given Proposition 1.

### 5.2 A Proof for a Restricted Parameter and Policy Set

#### Proposition 2.

*Under the conditions of Proposition 1, for any t* ≥ 0 *and i* ∈ *{*1, …, *n}*

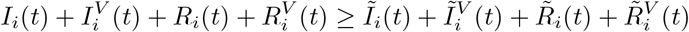

*and*

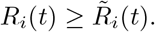

*Moreover, for any λ* ∈ [0, 1]

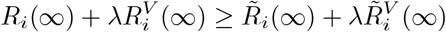

*and hence, the objective function is lower for Ũ, provided the conditions of Proposition 1 are met*.

**Proof:** Note that, by Proposition 1,

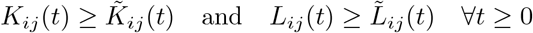

and hence, by Lemma A.2, for each *i* ∈ *{*1, …, *n}*

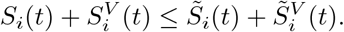

Combining this with the conservation of population equation, (13), shows that

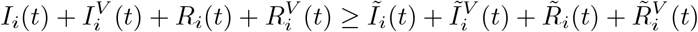

as required. Now, taking *t* → ∞ and noting that the infections tend to zero by Lemma B.4 gives

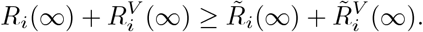

Moreover, by Lemma A.5, for any *t* ≥ 0 and *i* ∈ *{*1, …, *n}*

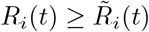

as required. Also, taking *t* → ∞ shows that

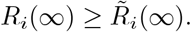

Thus, for any *λ* ∈ [0, 1]

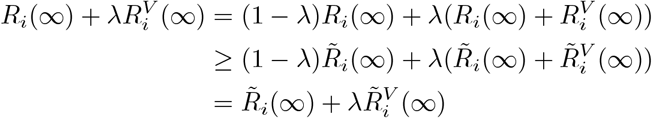

as required.

By summing the *i* inequalities at *t* = ∞ from Proposition 2 (and using *λ* = *κ*_*i*_), Theorem 1 holds under the additional conditions given in Proposition 1. Note that the closure of the set of parameters, initial conditions and vaccination policies which satisfy these conditions is the original set specified in Theorem 1. Thus, one can generalise the result with the help of the following proposition.

### 5.3 Continuous Dependence

#### Proposition 3.

*Define the set of functions*

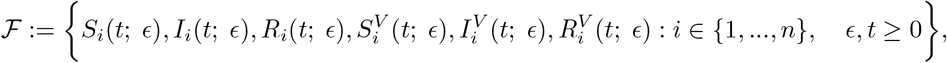

*where for each fixed ϵ, these functions solve the model equations with parameters*

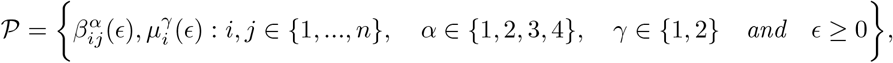

*initial conditions*

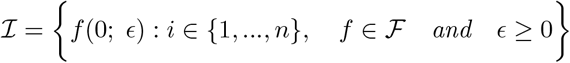

*and vaccination policy* ***U*** (*t*; *ϵ*). *Suppose that*

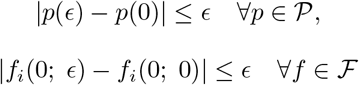

*and that*

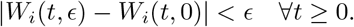

*Moreover, suppose that for each i* ∈ *{*1, …, *n} and ϵ* ≥ 0,

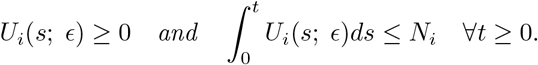

*Then, for each δ >* 0 *and each T >* 0 *there exists some η >* 0 *(that may depend on T and δ) such that*

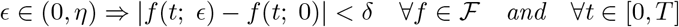

**Proof:** The proof is simple but algebraically dense and so is left to Lemma B.8 in the appendices.

This now allows a proof of Theorem 1 to be formed.

### 5.4 Theorem 1

#### Theorem 1.

*Suppose that* ***U***, ***Ũ*** ∈ *C. Suppose further that for each i* ∈ *{*1, …, *n} and t* ≥ 0

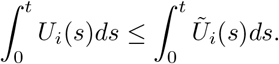

*Then*

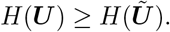

**Proof:** Define the parameters 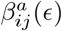 and 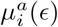 by

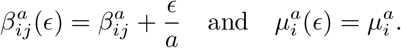

This means that, for any *ϵ >* 0, these parameters satisfy the conditions of Propositions 1 and 2. Define, for *ϵ <* 1, the initial conditions

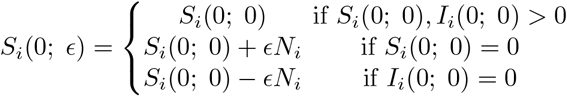

and

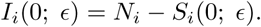

Then, the conditions of Propositions 1 and 2 are met by these initial conditions for any *ϵ >* 0.

Now, define the set of points

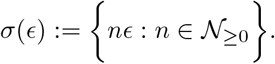

Then, define 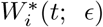 to be the first order approximation to the function 𝒲_*i*_(*t*; *ϵ*) := max(*W*_*i*_(*t*), *N*_*i*_ − *ϵ*) using the points of *σ*(*ϵ*). That is, for each *t* define

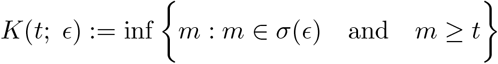

and

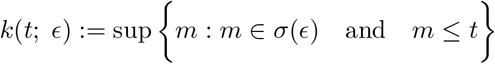

Note that, as *σ*(*ϵ*) is nowhere dense, one must have

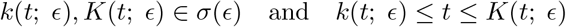

Then, define

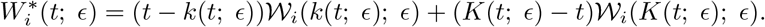

Thus, as *k* and *K* are constant on any interval not containing a point in *σ*(*ϵ*), 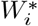 is linear on any interval not containing a point of *σ*(*ϵ*) and so its derivative is a step function.

Now, note that, for each *t*

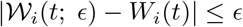

and, moreover,

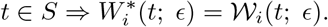

Also, as *U*_*i*_ is bounded, each *W*_*i*_ (and hence each 𝒲_*i*_) are Lipschitz continuous with some Lipschitz constant *L*. Moreover, each 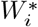 is continuous and is differentiable in each interval (*k*(*t*; *ϵ*), *K*(*t*; *ϵ*)) with a maximal (uniformly bounded) gradient of *U*_*i*_(*t*), meaning that 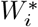 is also Lipschitz continuous with Lipschitz constant *L*.

It can now be shown that 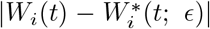 is uniformly bounded in *t*. For each *t* ≥ 0, one can find an element *s* ∈ *σ*(*ϵ*) such that |*t* − *s*| *< ϵ*. Then,

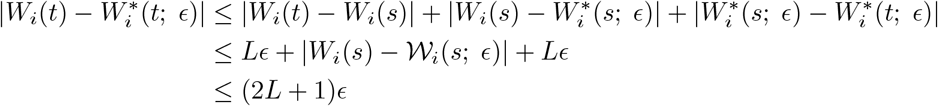

and so 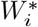 converges uniformly to *W*_*i*_. The same results hold for the analogously defined 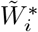. Then, note that, as 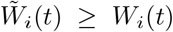, it must be that 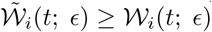. Thus, it follows that 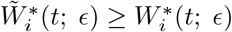.

This means that Proposition 2 can be used. Define using stars the variables that come from the ***U*** ^*^ and ***Ũ*** * policies. Then, from Proposition 2, for each *t* ≥ 0, *ϵ >* 0 and *i* ∈ *{*1, …, *n}*

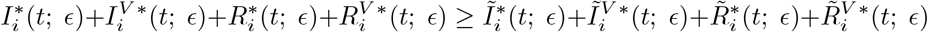

and

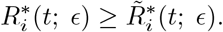

Then, taking *ϵ* → 0 and using Proposition 3 (noting that the perturbations to the parameters, initial conditions and vaccination policies are all bounded by a constant multiple of *ϵ*) shows that

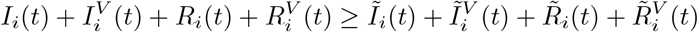

and

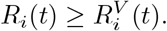

Then, the result follows using the same logic as in the proof of Proposition 2.

### 5.5 Theorem 2

#### Theorem 2.

*Suppose that B is differentiable, and that there is an optimal solution* ***U*** *to (6). Then, define the function*

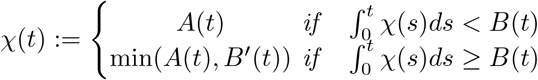

*and suppose that χ*(*t*) *exists and is bounded. Then, there exists an optimal solution* ***Ũ*** *to the problem (6) such that*

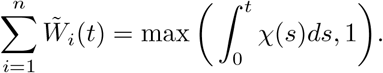

*Moreover, if χ*(*t*) *is continuous almost everywhere, there exists an optimal solution* ***Ũ*** *such that*

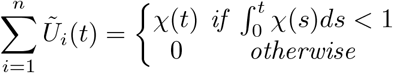

**Proof:** Suppose that ***U*** is an optimal vaccination policy. To begin, it will be shown that the total vaccination rate *χ* is indeed a maximal-effort vaccination policy (in the sense that, at each time *t*^*^, it is impossible to have given out more vaccines than a policy with total overall rate *χ*(*t*)).

**Claim:** 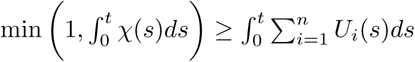 for all *t >* 0

**Proof:** Consider any time *t* ≥ 0 such that

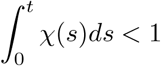

and define the set

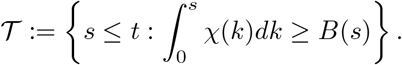

Suppose that 𝒯 = ∅. Then,

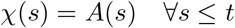

and so

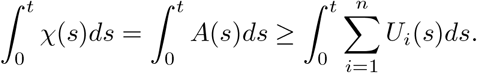

Moreover, suppose that 𝒯 ≠ ∅ and define

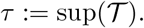

Then,

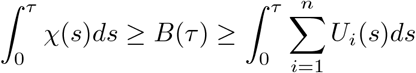

and

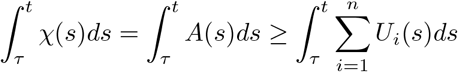

so that

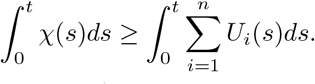

Thus, this holds in all cases for 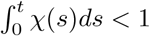. Finally, suppose that

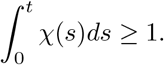

Then, one has

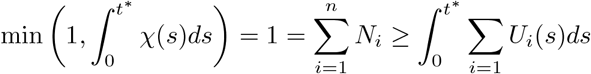

and so the claim is proved.

It is now important to show that *χ* gives a feasible vaccination rate. Note that *χ*(*t*) ≤ *A*(*t*) by definition.

**Claim:** 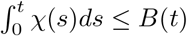 for all *t* ≥ 0.

**Proof:** Suppose, for a contradiction, that there exists a *t* such that

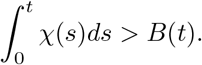

Then, define

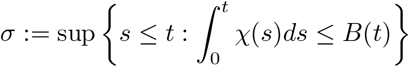

which must exist (as 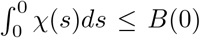) and satisfy *σ < t*, by continuity of 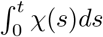 and *B*(*t*). Note that

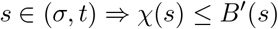

and so

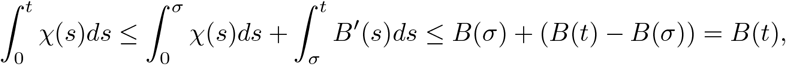

which is a contradiction. Thus,

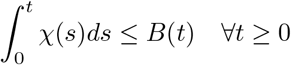

as required.

Now, one can create a new optimal vaccination policy with total rate given by *χ*. Define

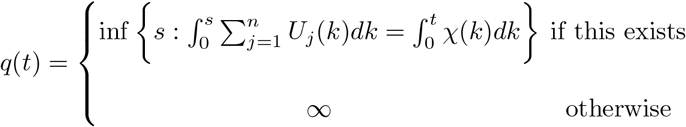

so that *q*(*t*) represents the earliest time at which *χ*(*t*) vaccines were administered by the ***U*** policy. By continuity of the integral, this means that

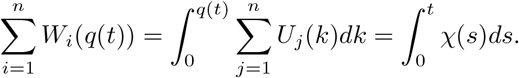

Define further

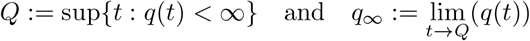

so that *Q* is the earliest time at which all of the vaccines given out by the ***U*** policy could have been administered. Note that both *Q* and *q*_∞_ may be infinite. By taking the limit *t* → *Q*, and noting the left hand side is bounded by 1,

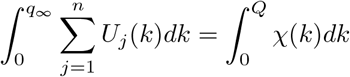

Then, the integral of the new vaccination policy, 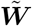 is given by

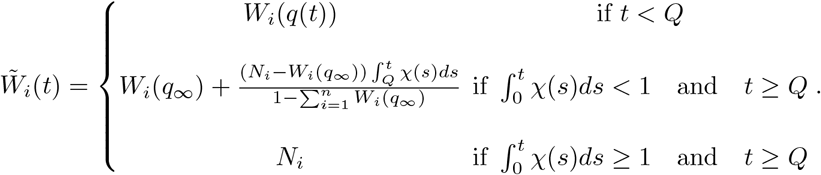

This is well-defined as

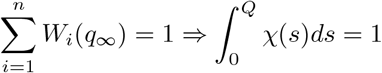

and so, in this case, the second part of the definition of *χ* is never used. It is important to establish for feasibility that each *W*_*i*_ is bounded by *N*_*i*_.

**Claim:** 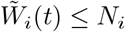 for all *t* ≥ 0 and all *i* ∈ *{*1, …, *n}*.

**Proof:** If *t < Q*, then *W*_*i*_(*q*(*t*)) ≤ *N*_*i*_ for all *t < Q* by feasibility of ***U***. Otherwise, if *t* ≥ *Q* and 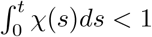, then one has

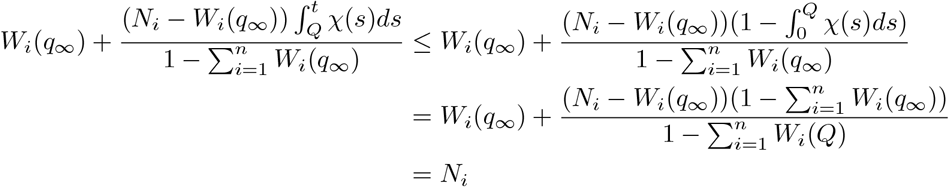

while if 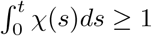 then the result is immediate.

The optimisation problem is framed in terms of ***U*** rather than ***W***, and so it is important to show that there is some ***Ũ*** that integrates to 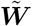. One can do this by proving the Lipschitz continuity of 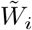 for each *i*.

**Claim:** 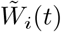 is Lipschitz continuous for each *i* ∈ *{*1, …, *n}*

**Proof:** Note that for *s, t < Q*, if *M* is a bound for *χ* (which is assumed to exist)

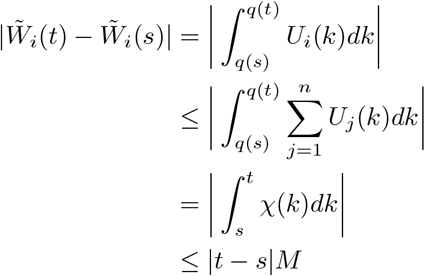

Moreover, if *s, t > Q* and 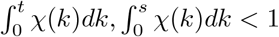, then

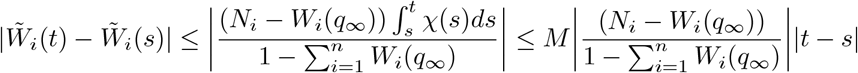

and if *s, t > Q* and 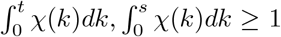, then 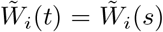. The intermediate cases (where *s* and *t* correspond to different cases in the definition of *χ*) can be proved by combining these bounds.

This means that (for each *i*) there exists a Lebesgue integrable function *Ũ*_*i*_(*t*) such that

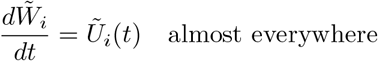

and, for all *t* ≥ 0

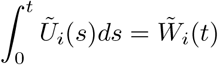

A proof of this (for the broader class of absolutely continuous functions) can be found in Bárcenas (2000). One can set *Ũ*_*i*_(*t*) to be zero for any *t* such that 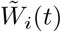 is not differentiable. Thus, noting that, where it is differentiable, the derivative of 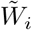 is bounded by its Lipschitz constant, *Ũ*_*i*_(*t*) is bounded as required.

Note that, in all cases (as 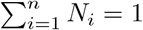)

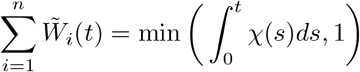

and so 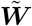 does correspond to a maximal vaccination rate. If *χ*(*t*) is continuous almost everywhere, then one can differentiate this relationship at *t* where each 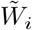 is differentiable and *χ* is continuous to show that 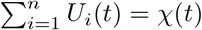. The complement of this set must have zero measure (as it is the finite union of zero measure sets), and so, in this case, one can change the values of each *U*_*i*_(*t*) so that 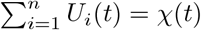 everywhere without changing the value of ***W***.

**Claim:** 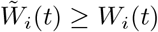 for all *i* ∈ *{*1, …, *n}* and *t* ≥ 0

**Proof:** Note that, by maximality of *χ*, for *t < Q*,

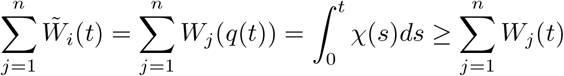

If *q*(*t*) ≥ *t*, then *W*_*i*_(*q*(*t*)) ≥ *W*_*i*_(*t*) for each *i*. If *q*(*t*) *< t*, then it is necessary that *W*_*i*_(*q*(*t*)) = *W*_*i*_(*t*) for each *i* as *W*_*i*_ is non-decreasing. Thus, *W*_*i*_(*q*(*t*)) ≥ *W*_*i*_(*t*) for all *i* and for all *t < Q*.

If *t > Q* and 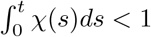, then

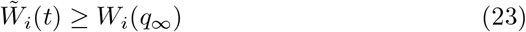

Now, by definition of *Q*, it is necessary that

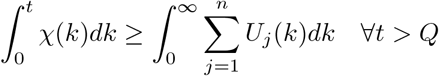

as otherwise, there must exist some *t > Q* and some *s <* ∞ such that

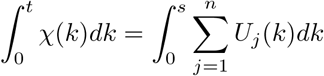

which means that *q*(*t*) *<* ∞. Thus, by continuity, for all *τ* ∈ (0, *t*), there exists some *s* such that

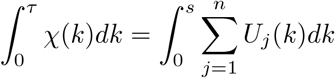

which means *Q* ≥ *t*, which is a contradiction.

Thus, by taking *t* → *Q*,

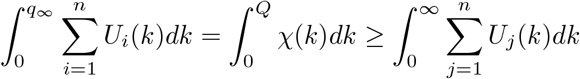

and so

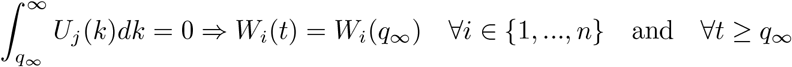

Thus, using (23),

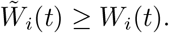

Finally, if *t > Q* and 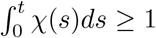, then 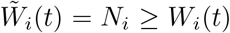. Thus, for all *t* and *i*,

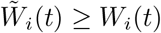

as required.

Thus, by Theorem 1, it is necessary that

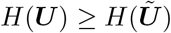

and hence, by the optimality of ***U***, ***Ũ*** is optimal as required.

### 5.6 Theorem 3

#### Theorem 3.

*Under the assumptions of Theorem 2, consider a modified objective function* ℋ *given by*

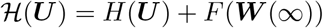

*for any function F. Then, with χ defined to be the maximal vaccination effort as in Theorem 2, there exists an optimal solution* ***Ũ*** *such that, for some τ* ≥ 0

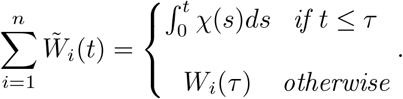

*Moreover, if χ is continuous almost everywhere, then there is an optimal solution* ***Ũ*** *such that*

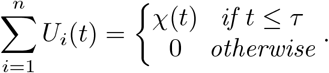

**Proof:** This follows directly from the proof of Theorem 2. One can again define ***Ũ*** in the interval (0, *Q*) (where *Q* is defined in the proof of Theorem 2) such that

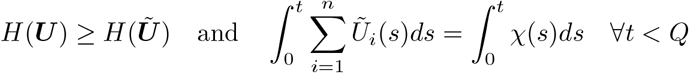

with the only difference being that now

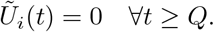

Thus, as shown in the proof of Theorem 2,

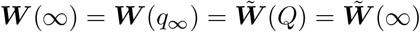

and so

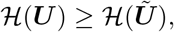

which means ***Ũ*** is optimal as required.

## 6 Limitations of Theorem 1

It is helpful to consider the limitations of Theorem 1, as it does not prove that every conceivable cost function is non-increasing in vaccination effort. This will be illustrated through some examples based on theoretical COVID-19 outbreaks in the United Kingdom.

Using the work of Prem et al (2017), one can split the UK into 16 age-groups (comprising five year intervals from 0 to 75 and a group for those aged 75+) which mix heterogeneously. The contact matrices estimated in Prem et al (2017) allow for the construction of a matrix ***β***^*^, which will be proportional to each of the matrices ***β***^*α*^ in the model.

As illustrated in Liu et al (2020), estimation of the basic reproduction number *R*_0_ for COVID-19 is complicated, and a wide range of estimates have been produced. For the examples in this paper, a reproduction number of 4 will be used, meaning that *β*^1^ will be scaled so that the largest eigenvalue of the matrix given by

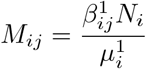

is equal to 4. Note that the population of each group ***N*** - normalised to have total sum 1 - is taken from uni (2019). Moreover, based on the estimates in Ram and Schaposnik (2021), the value of 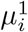 and, in the first example, 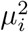 will be set equal to 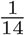.

To model the effectiveness of vaccination, the estimates of Dean and Halloran (2022) will be used so that *β*^2^ = 0.77*β*^1^, (modelling the reduction in infectiousness), *β*^3^ = 0.3*β*^1^ (modelling the reduction in susceptibility) and *β*^4^ = 0.77 × 0.3 × *β*^1^ (assuming these effects are independent). Finally, the initial conditions used are *S*_*i*_(0) = (1 − 10^−4^)*N*_*i*_ and *I*_*i*_(0) = (10^−4^)*N*_*i*_ for each *i*, modelling a case where 0.01% of the population is initially infected. It should be emphasised however, that this model has purely been made for illustrative purposes and substantially more detailed fitting analysis would be required to use it for forecasting COVID-19 in the UK.

In both the subsequent examples, it will be assumed that 0.5% of the population is vaccinated homogeneously each day in the vaccination case. This will be compared to a case with no vaccination.

### 6.1 Infections Are Not Decreasing For All Time

While the overall number of infections will decrease as vaccination effort increases, the infections at a particular point in time will not. Figure 2 shows that the effect of vaccination is both to reduce, but also delay the peak of the infections. This is an important consideration when deciding vaccination policy, as increasing infections at a time in the year when hospitals are under more pressure could have negative consequences, and so it is important not to simply assume that vaccination will reduce all infections at all times.

**Fig. 2.**
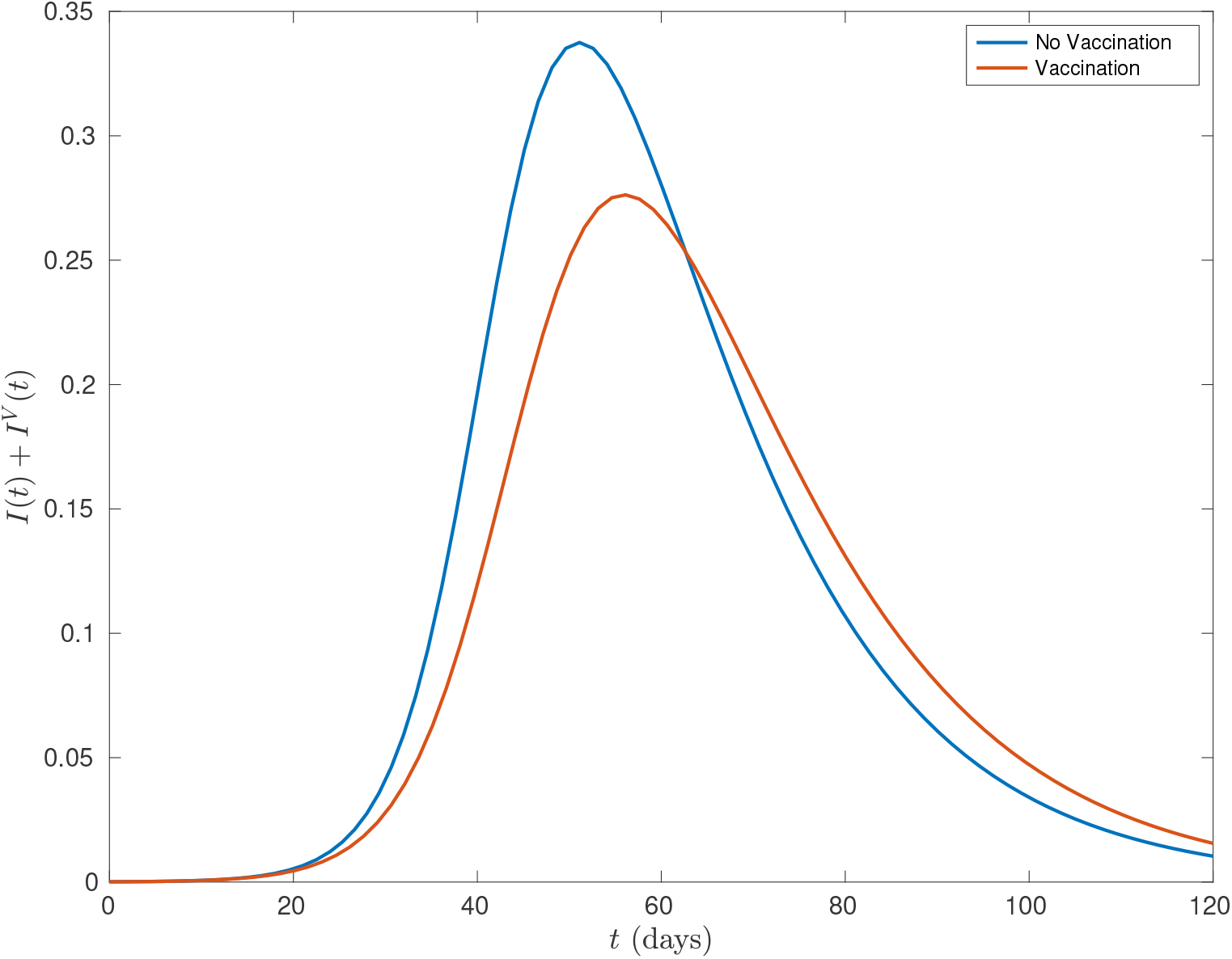
A comparison of the total infections over time for a simulated COVID-19 epidemic in the UK, depending on whether a uniform vaccination strategy of constant rate is used.

### 6.2 Deaths Are Not Decreasing For All Time

Perhaps most surprisingly, the total deaths in the epidemic may at some finite times (although not at *t* = ∞) be higher when vaccination occurs, at least under the assumptions of the SIR model. This is a rarer phenomenon, but is possible if vaccination increases the recovery rate as well as decreasing infectiousness.

For illustrative purposes, suppose that vaccination doubles the recovery rate (so that 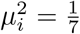) and has no effect on mortality rates. Then, using Bonanad et al (2020) to get age-dependent mortality rates for COVID-19, Figure 3 shows that initially, the number of deaths is higher in the case of vaccination. This occurs because the higher value of *μ*^2^ means that vaccinated people move more quickly to the *R*^*V*^ compartment than their unvaccinated counterparts and so, while they will infect fewer people, when the number of infections is comparable in the early epidemic, this means that more people will die. Of course, this is not a realistic reflection of the course of an epidemic - the reason for *μ*^2^ being higher is that vaccinated people are likely to get *less* ill rather than dying more quickly - but it illustrates that the model equations can have interesting, and unexpected properties.

**Fig. 3.**
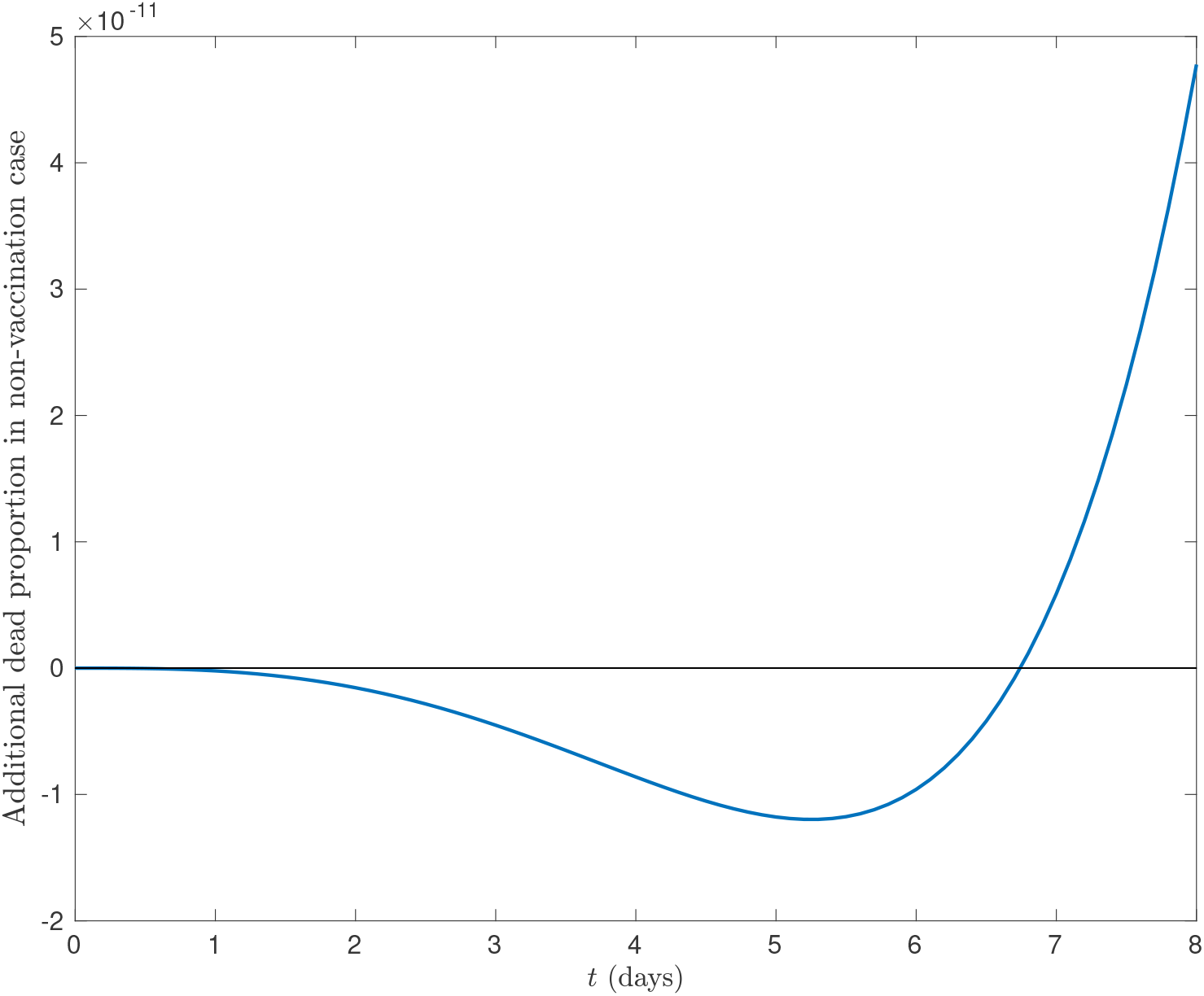
The difference between proportion of the population that has died by each time *t* in the case of vaccination and non-vaccination. Positive values indicate that the deaths are higher in the non-vaccination case.

## 7 Discussion

It is comforting that the multi-group SIR model does indeed satisfy the condition that the final numbers of infections and deaths are non-increasing in vaccination effort. This shows the importance of ensuring that vaccinations are available as early as possible in a disease outbreak. To achieve this, it is important that good plans for vaccine roll-out and supply chains are available in advance of them being needed to ensure that maximum benefit from the vaccination program is obtained.

However, there are limitations to these results. Indeed, while the final numbers of infections and deaths are guaranteed to decrease, this is not necessarily true at a given finite time. In particular, vaccination can move the peak of the epidemic, and so it is important to consider the consequences of this, particularly if only a small number of lives are saved by vaccination.

Moreover, while this has not been discussed in this paper, it is also important to emphasise that these results only apply if vaccine efficacy does not decay over time. Indeed, if vaccination efficacy does decay significantly, then vaccinating the most vulnerable groups in a population very early may be worse than vaccinating them later, unless booster jabs are available. If the main epidemic occurs long after the vulnerable have been vaccinated, their immunity may have worn off significantly by the time that the majority of disease exposure occurs. Thus, in this case a more detailed analysis would be needed to determine the optimal vaccination rate.

It seems to the authors that future models for optimal vaccination should use the unvaccinated-only model used in in this paper, where vaccines are distributed randomly to the unvaccinated, rather than the entire population. While for small total vaccination numbers, the difference between the two models is small, when a significant proportion of the population is vaccinated, there can be large differences between the optimal policies, as was shown through the example in Section 2. Moreover, it appears to better reflect the vaccine uptake from the COVID-19 pandemic, as was illustrated in Figure 1.

Of course, this modified model is slightly more complicated, and care needs to be taken to avoid numerical instabilities arising from the removable singularity in the 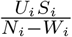 term when *W*_*i*_ → *N*_*i*_. However, it has been shown that many of the standard properties of SIR models, and indeed the results of this paper, still hold for this model, and so these extra technical difficulties appear to be a small price to pay for the significantly increased accuracy and potentially large difference between the optimal solutions for the two models.

The results of this paper could be extended to cover a wider range of disease models that are currently being used in the literature. In particular, the next step could be to prove the results for SEIR (Susceptible-Exposed-Infected-Recovered) models, and indeed models with multiple exposed compartments for each subgroup. This would help to build a general mathematical theory of maximal-effort vaccination that would provide evidence for the reliability of contemporary epidemiological modelling.

## 8 Conclusion

The results of this paper are summarised below:

- Vaccinating at maximal effort is optimal for a multi-group SIR model with non-decaying vaccination efficacy.
- The “random vaccination model” found in the literature may overestimate the decay of vaccination uptake in a population and so the “unvaccinated-only model” presented here should be considered for finding optimal vaccination policies.
- While vaccinating at maximal effort gives optimality, there can be finite times at which, according to the SIR model, infections or deaths are higher if vaccination has occurred.

## Data Availability

Links are provided in the manuscript to all data used in the present work.

https://ourworldindata.org/coronavirus

## Declarations

This work was funded by a DTP Studentship awarded to Matthew Penn by the Engineering and Physical Sciences Research Council (EPSRC) through the University of Oxford (https://www.ukri.org/councils/epsrc/career-and-skills-development/studentships/doctoral-training-partnerships/). For the purpose of Open Access, the author has applied a CC BY public copyright licence to any Author Accepted Manuscript (AAM) version arising from this submission. The funders played no specific role in any aspect of this work.

The authors declare no competing interests.

## Data Availability

As cited in the text, the data used to create Figure 1 is available from https://github.com/owid/covid-19-data/blob/master/public/data/vaccinations/vaccinations-by-age-group.csv. The data used to create Figure 2 is available from https://journals.plos.org/ploscompbiol/article?id=10.1371/journal.pcbi.1005697 and https://population.un.org/wpp/Download/Standard/Population/. The additional data used to create Figure 3 is available from https://www.sciencedirect.com/science/article/pii/S1525861020304412.

## Acknowledgements

The authors would like to thank Cameron Simmons, Joseph Penn and Grace Penn for their invaluable proof-reading work.

## A Supplementary Lemmas For Proposition 1 and 2 and Theorem 2

For the proofs of these lemmas, it is helpful to recall the following definitions of the following variables, which will be extensively used.

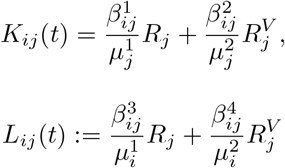

and

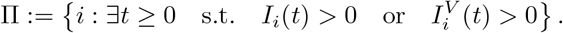

Moreover, note that, under the assumptions of Proposition 1 and 2, each *U*_*i*_(*t*) is a step function and is therefore piecewise smooth in each bounded interval. Thus, in particular, the derivatives of each of the model variables (and indeed, the derivative of *W*_*i*_(*t*)) are piecewise continuous in each bounded interval, meaning that each of the model variables is piecewise continuously differentiable in each bounded interval. This means that integration by parts can be performed (in a bounded interval), as will be done extensively throughout the proofs of these lemmas.

### A.1 Lemma A.1

#### Lemma A.1.

*Suppose that f* (*t*) *is a non-increasing, non-negative, continuous and piecewise continuously differentiable function and that the continuous and piecewise continuously differentiable functions g*(*t*) *and h*(*t*) *satisfy g*(0) = *h*(0) *and g*(*t*) ≤ *h*(*t*) *for all t* ≥ 0. *Then*,

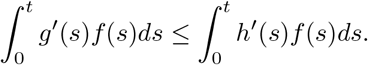

**Proof:** This follows from integrating by parts:

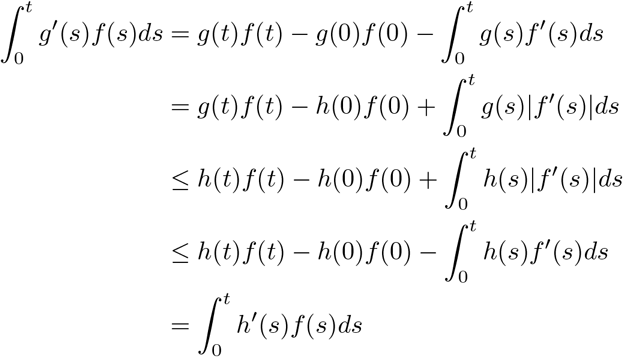

as required.

### A.2 Lemma A.2

#### Lemma A.2.

*Suppose that*

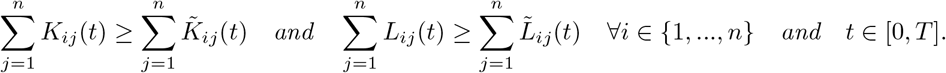

*Then*,

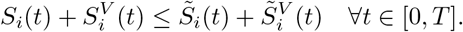

**Proof:** To reduce notation in this proof, define

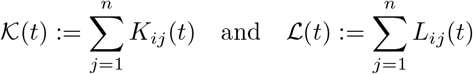

Note that

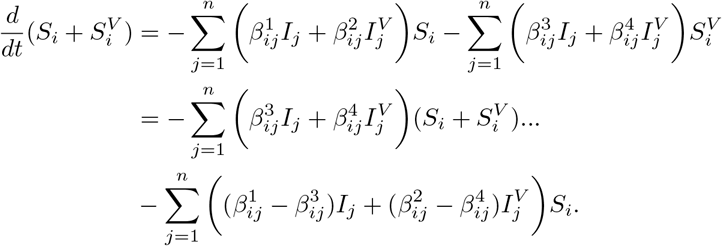

Thus,

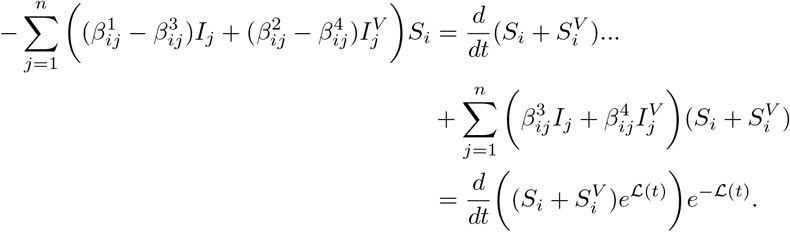

This means that

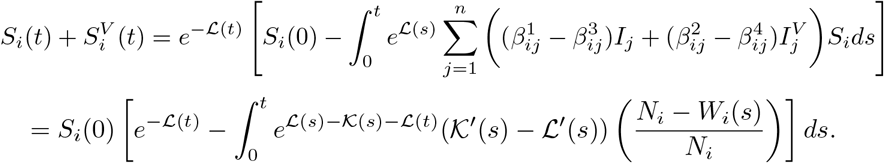

Now, one can see that, as 0 ≤ *W*_*i*_(*s*) ≤ *N*_*i*_,

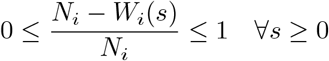

and hence

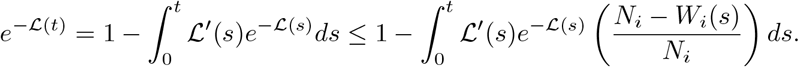

Now, this means that

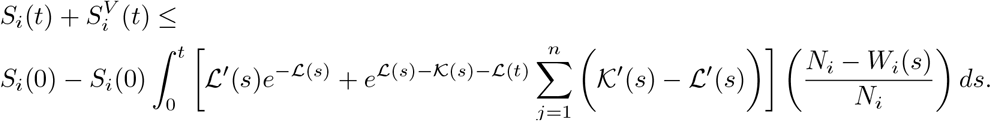

This allows the use of Lemma A.1. Firstly, note that, as 𝒦′(*s*) ≥ ℒ′(*s*) ≥ 0 and 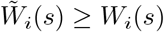, one has

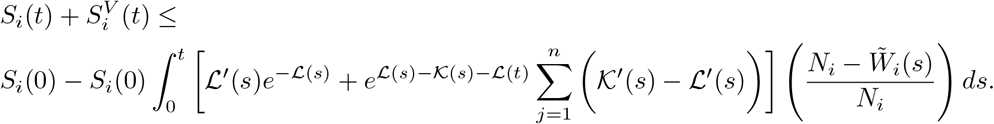

Moreover,

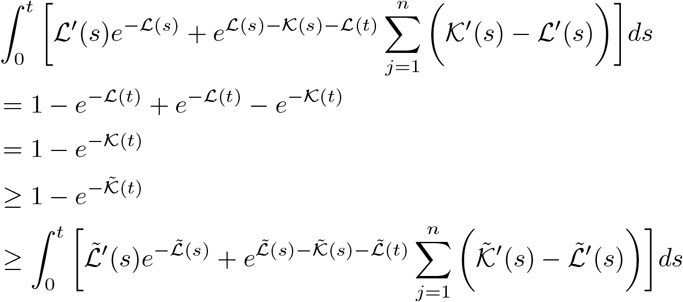

and *N*_*i*_ − *W*_*i*_(*s*) is non-increasing in *s*. Thus, by Lemma A.1, with

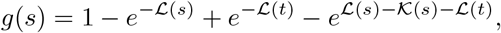

*h*(*s*) defined as the tilde version of *g*(*s*), and *f* (*s*) := *N*_*i*_ − *W*_*i*_(*s*), one has

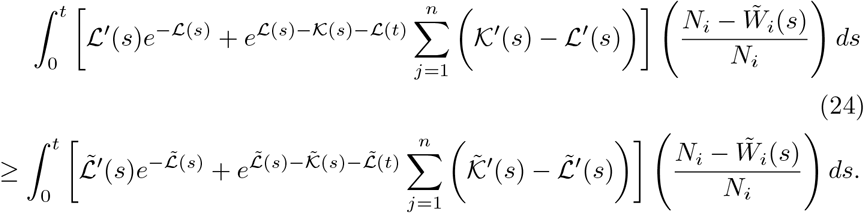

Thus, (as this integral is multiplied by -1 in (24)), combining this with (24) gives

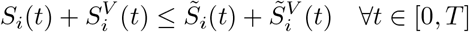

as required

### A.3 Lemma A.3

#### Lemma A.3.

*Suppose that*

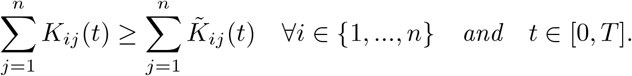

*Then*

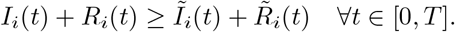

To begin, one can write the equation for *S*_*i*_ as

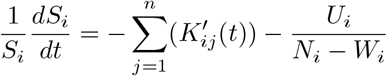

and hence, integrating

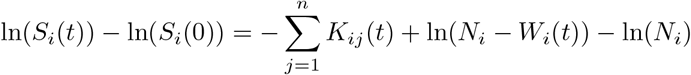

which implies

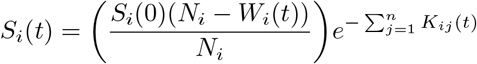

Using this result shows that

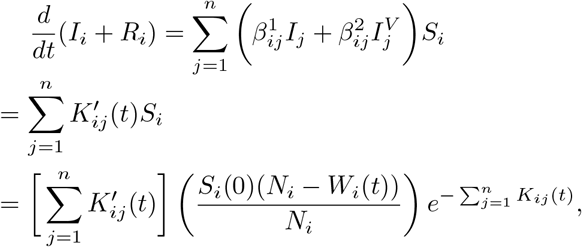

Thus,

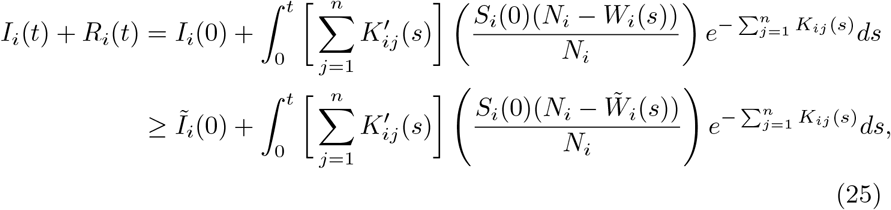

using the fact that the initial conditions are the same in both cases and that 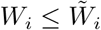. Now, one can use the results of Lemma A.1 with

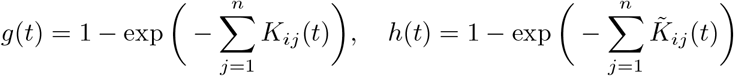

and 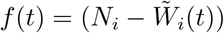, noting that

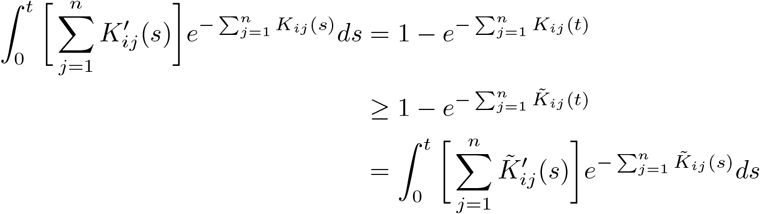

and that 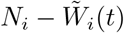 is non-increasing. Thus,

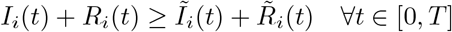

as required.

### A.4 Lemma A.4

#### Lemma A.4.

*Suppose that*

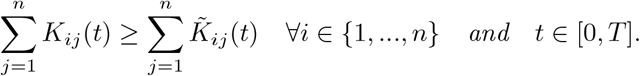

*Then*,

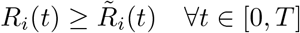

**Proof:** The result of Lemma A.3 can be written as

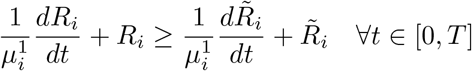

which implies

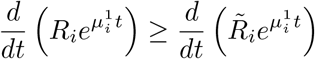

and hence, after integrating and cancelling exponentials, one finds

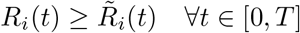

as required.

### A.5 Lemma A.5

#### Lemma A.5.

*Suppose that*

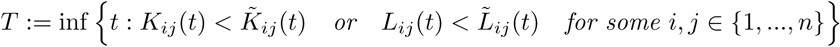

*exists. Then, for some b* ∈ *{*1, …, *n}, and some real constants κ and η*,

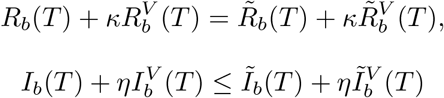

*and*

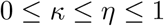

**Proof:** Suppose that *T* exists. Then, by continuity, there exists some *a* and *b* such that 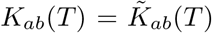 or 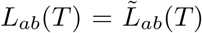. These can be rearranged to give, respectively,

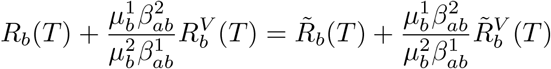

or

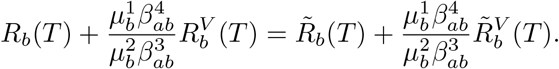

This can be written as

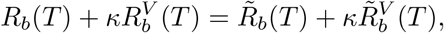

where, by the inequality constraints on the 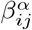 and 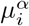

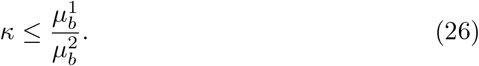

Moreover, note that

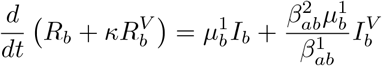

is a continuous function. Thus, if

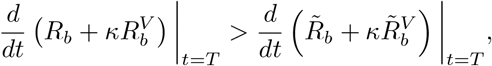

then there exists some *τ >* 0 such that

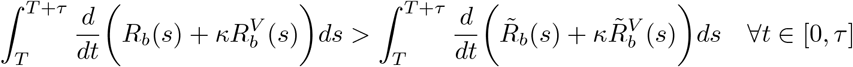

and hence, in particular

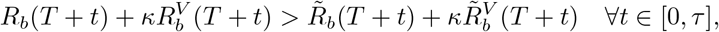

Thus, it is necessary that there is some *b* such that

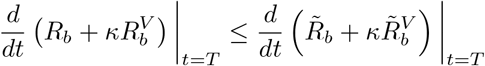

so

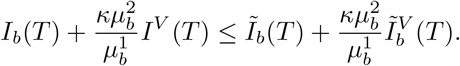

This can be written as

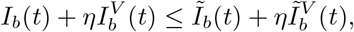

where, by (26), the fact that 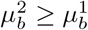, and the non-negativity of all parameters,

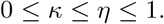

as required

### A.6 Lemma A.6

For the purposes of this lemma, it is helpful to recall the inequality system (7)-(13).

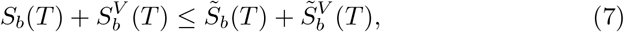

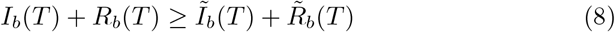

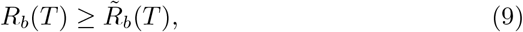

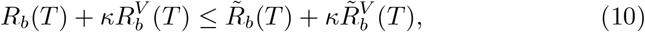

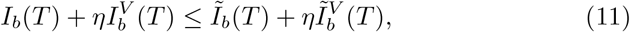

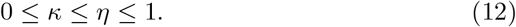

and

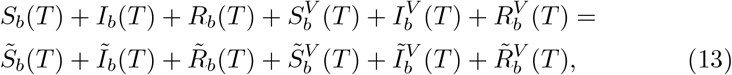

#### Lemma A.6.

*Suppose that the system (7) - (13) holds for some b* ∈ *{*1, …, *n} and some T* ≥ 0. *Then*,

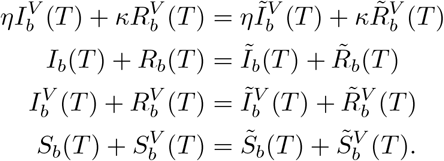

**Proof:** To begin, note that adding inequalities (7), (10) and (11) gives

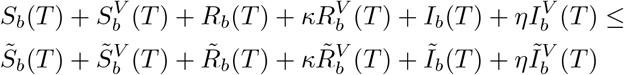

and then, using (13) shows that

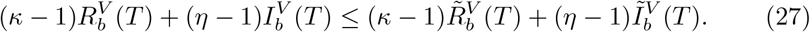

Moreover, adding (10) and (11) shows that

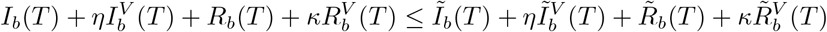

and then, using (8) shows that

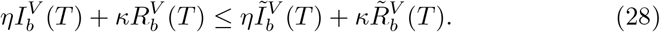

Now, from the inequality (10) combined with the inequality (9), it must be the case that

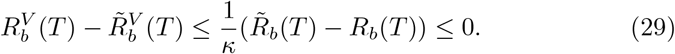

Define

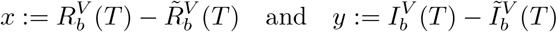

so that the system given by (12), (27), (28) and (29) reduces to

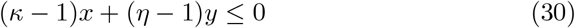

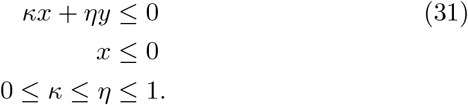

Note first that *x* = 0 implies that *y* = 0 as *η* and (*η* − 1) have different signs. Thus, in this case, the inequalities (30) and (31) are in fact equalities.

Suppose instead that *x* ≠ 0 (so *x <* 0). The first two of these inequalities can be rearranged (noting the signs of the denominators) to give

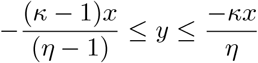

and so, as −*x >* 0,

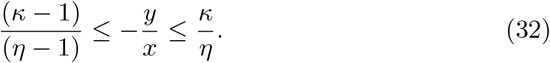

However, note that

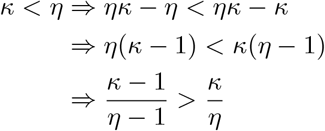

and hence, as *κ* ≤ *η*, for there to be solutions to the inequality (32), it is necessary that

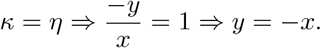

This means that the inequalities (30) and (31) are satisfied to equality in this and hence, from before, all cases. Thus, it is necessary that

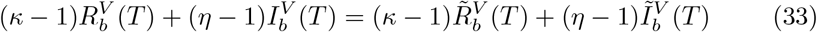

and

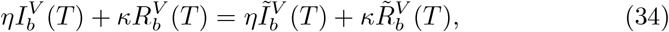

which is the first required equality. Thus, one can once again add the inequalities (10) and (11) to give

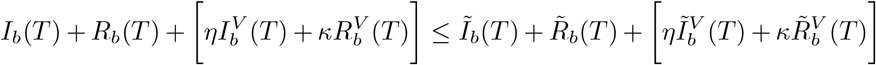

and so

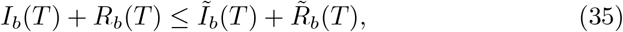

which, combined with (8), shows that

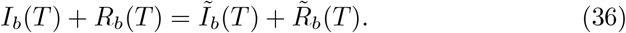

Moreover, one can subtract (33) from (34) to get

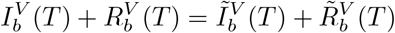

and then, using (13) alongside (35) and (36) shows

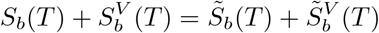

as required.

### A.7 Lemma A.7

Note that for this lemma, it will be assumed that each 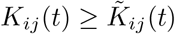, rather than the inequality simply holding for their sums as before.

#### Lemma A.7.

*Under the assumptions of Proposition 1, suppose that the system of inequalities (7) - (13) holds for some b* ∈ {1, …, *n*} *and some T >* 0. *Suppose further that*

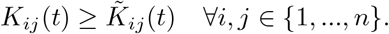

*Then*,

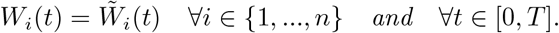

**Proof:** By Lemma A.6, the system (14) - (16) must hold for *b*. Now, Equation (25) in the proof of Lemma A.3 shows that

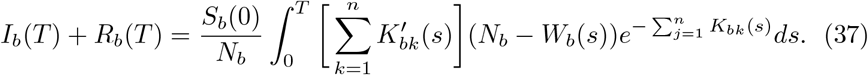

Now, the equality (14) shows

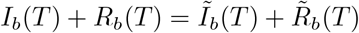

and hence, after cancelling the non-zero *S*_*b*_(0) and *N*_*b*_ terms, (37) (and its tilde equivalent) shows that

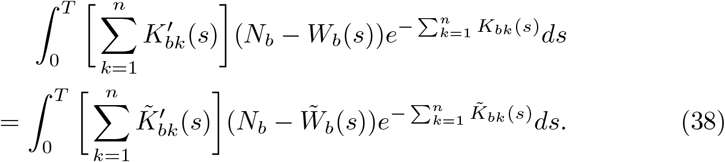

Note that, from Lemma B.6, as Π = *{*1,, *n}*

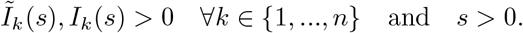

Thus,

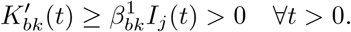

In particular,

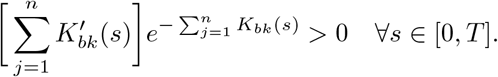

Moreover, by continuity of 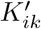 (as continuous functions attain their bounds on closed intervals), there exists some *m >* 0 such that

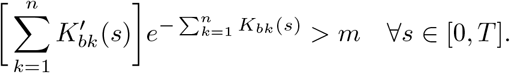

Hence, as 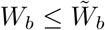

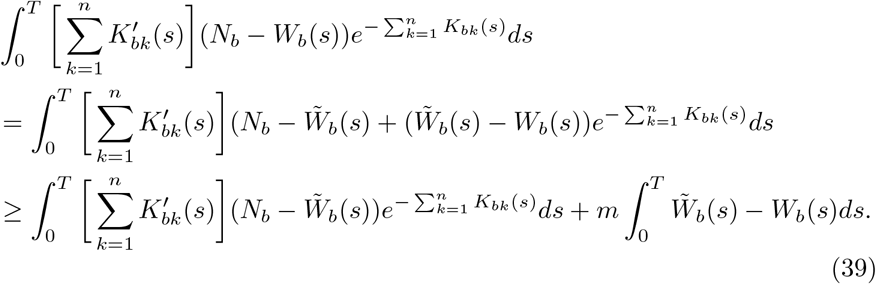

Finally, as 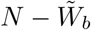 is decreasing and for any *t* ∈ [0, *T*],

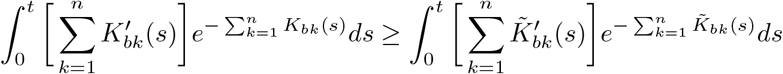

one has, by Lemma A.1, setting

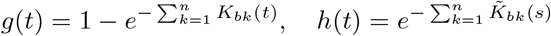

and 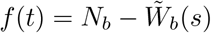,

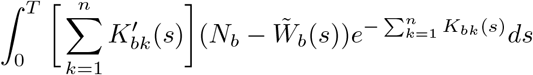

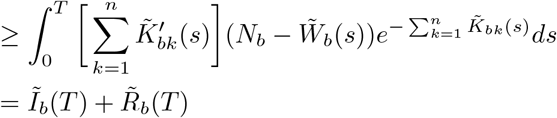

and so, combining this with (39),

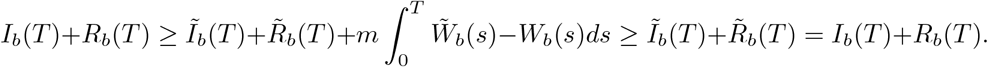

Hence,

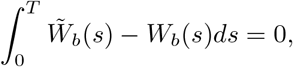

which by continuity means

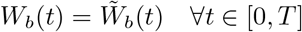

Now, moreover, substituting this back into the equality given in (38) shows that

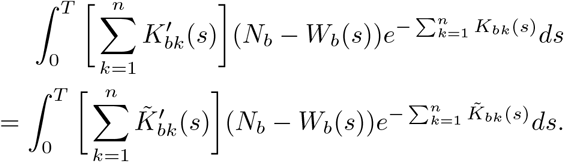

Hence, integrating by parts, this shows that

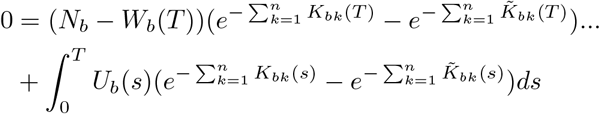

Now,

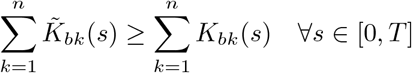

and so, for equality, it is necessary that

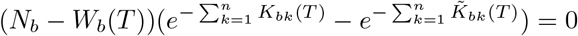

Thus, as it is assumed that *W*_*b*_(*t*) *< N*_*b*_ for all *t* ≥ 0,

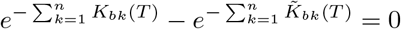

and hence, as 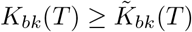 for all *k* ∈ *{*1,, *n}*,

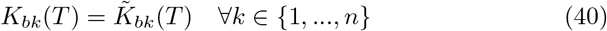

Now, suppose that 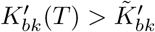 for some *k*. Then, by continuity and the fact that *T >* 0, it is necessary that there is some *τ* ∈ (0, *T*) such that

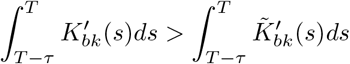

which means that

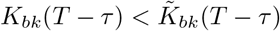

which is a contradiction to the definition of *T*. Thus, it is necessary that

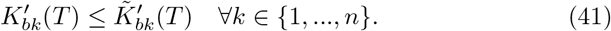

Dividing (40) by 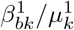 and (41) by 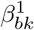 shows that the inequality system (7) - (13) holds for each *k* (as Lemmas A.2 - A.4 hold for any group) and so, following Lemma A.6 and the previous work of this proof, it is necessary that

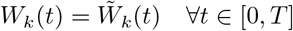

This holds for each *k* and hence the proof is complete.

### A.8 Lemma A.8

#### Lemma A.8.

*Define functions* 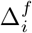 *to be*

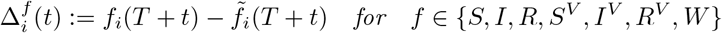

*and suppose that*

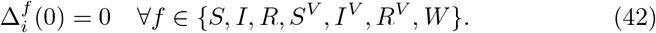

*Suppose further that the U*_*i*_(*t*) *are right-continuous step functions. Then, for t* ∈ [0, *δ*] *in the limit δ* → 0, *and for any x, y* ∈ ℜ

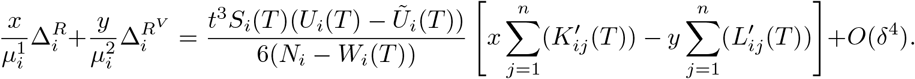

**Proof:** As the *U*_*i*_(*t*) are step functions, for sufficiently small *δ*, they are constant on the interval [*T, T* + *δ*], so this will be assumed. Note that, for any *i* ∈ *{*1, …, *n}* and any *t* ≥ 0

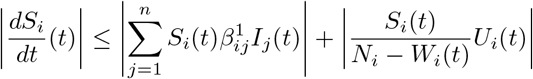

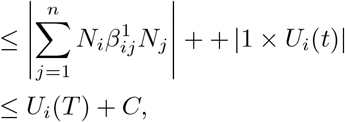

where the constant term, *C*, is independent of *t* and the vaccination policy. Note the second line follows from the fact that, as *W*_*i*_(*t*) *< N*_*i*_,

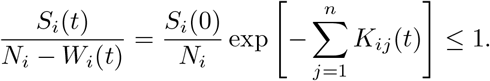

Similarly, one can show (by increasing the constant *C* if necessary) that

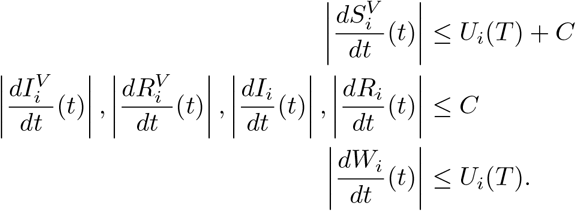

Then, for *t* ∈ (0, *δ*) and *f* ∈ *{S, I, R, S*^*V*^, *I*^*V*^, *R*^*V*^, *W}*

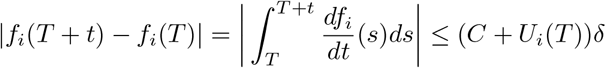

so that, in particular

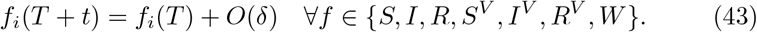

Now,

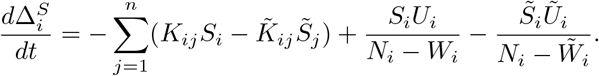

Using (42) and (43), this equation linearises to

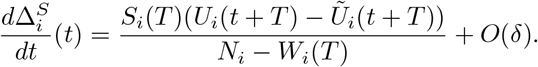

Noting that

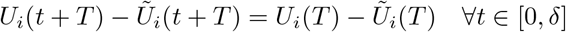

this means that

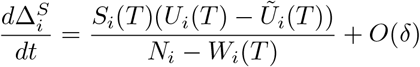

and so (for *t < δ*)

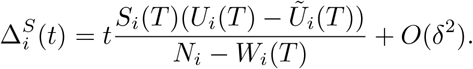

Now, one can linearise the equation for 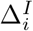. Note that

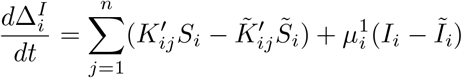

and so, with

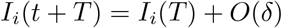

and similar expressions for other variables,

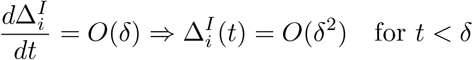

Now, one can linearise in a different way. Note that

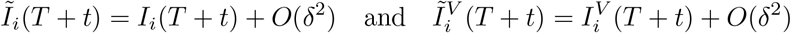

so

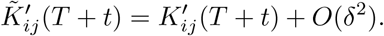

Thus,

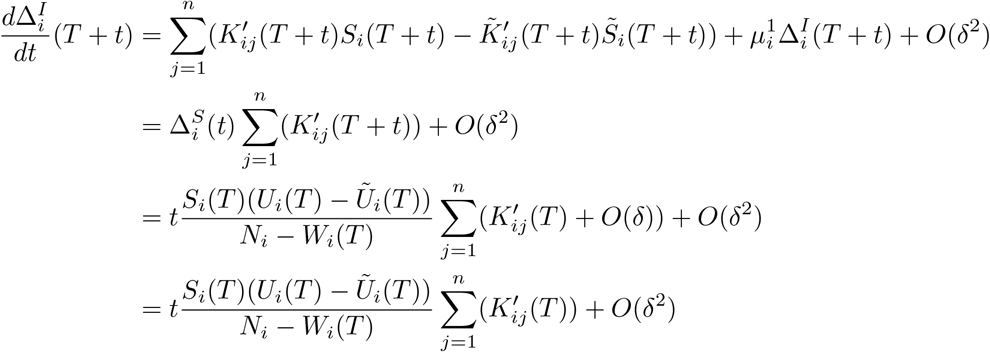

and hence

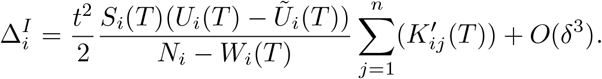

Thus,

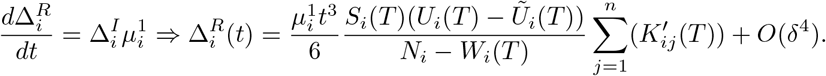

Now, note that

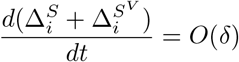

as this derivative has no explicit dependence on *U*. Thus, in particular,

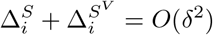

and so

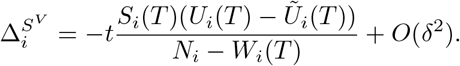

Then, as before (as the equation for 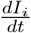 is the same as that for 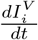, but with 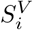 instead of *S*_*i*_, 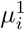 instead of 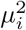 and *K*_*ij*_ instead of *L*_*ij*_)

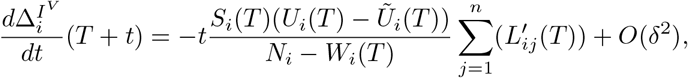

which means

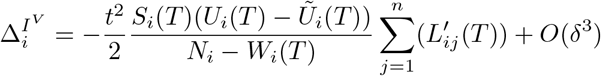

and hence

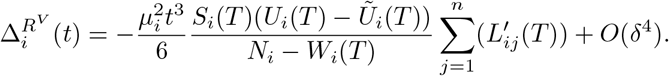

Thus,

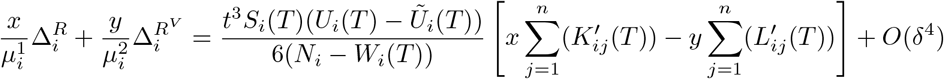

as required.

### A.9 Lemma A.9

#### Lemma A.9.

*Suppose that*

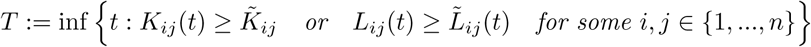

*exists. Define functions* 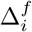 *to be*

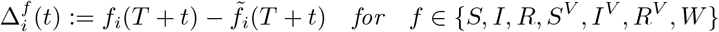

*and suppose that*

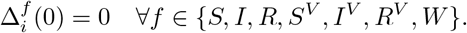

*Suppose further that the U*_*i*_(*t*) *are right-continuous step functions*, Π = *{*1, …, *n} and that*

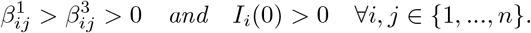

*Then*,

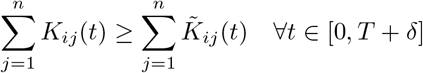

*and*

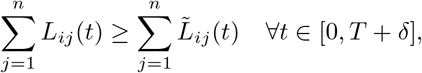

*for sufficiently small δ*.

**Proof:** By Lemma A.8, with 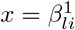 and 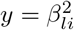 for some *l* ∈ *{*1, …, *n}*

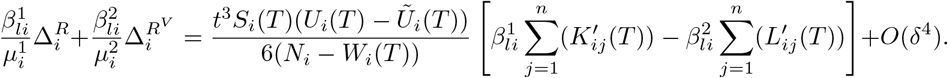

Now, as 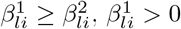 and 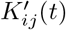 and 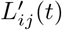 are non-negative

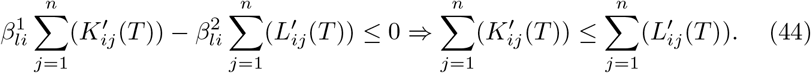

Noting that

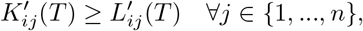

(44) requires

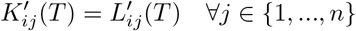

which, from the definitions of *K*′ and *L*′ requires

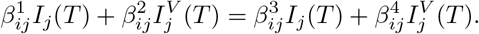

Thus, as *I*_*j*_(*T*) *>* 0 (as Π ∈ *{*1, …, *n}*) and 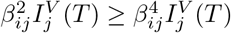, it is necessary that

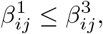

which is a contradiction. Thus,

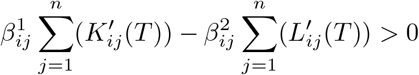

which means

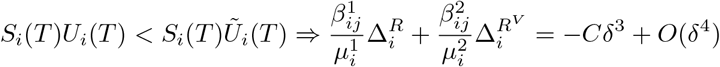

for some positive constant *C*. Now, if

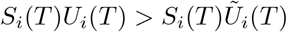

then, necessarily, *U*_*i*_(*T*) *> Ũ*_*i*_(*T*). Thus, as 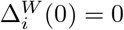, one will have

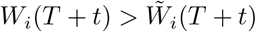

for sufficiently small *t*, which is a contradiction. Moreover, if

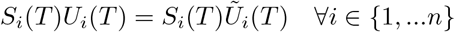

then the vaccination policies are the same in the interval [*T, T* + *δ*], as for each *i*, either *S*_*i*_(*T*) = 0 (in which case there is no more vaccination in group *i* so *U*_*i*_(*T*) = *Ũ*_*i*_(*T*) = 0) or *U*_*i*_(*T*) = *Ũ*_*i*_(*T*). Thus, the disease trajectories are the same, which contradicts the definition of *T*, as then 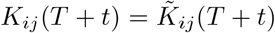 and 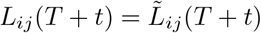 for all *t* ∈ [0, *δ*].

Now, note that

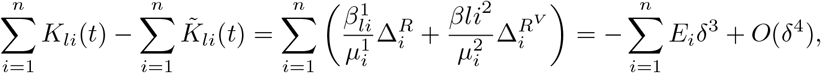

where *E*_*i*_ *>* 0 if *U*_*i*_(*T*) *< Ũ*_*i*_(*T*) and *E*_*i*_ = 0 otherwise. Thus, in particular

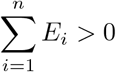

and hence

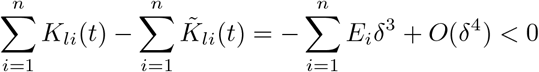

for sufficiently small *δ*. Thus,

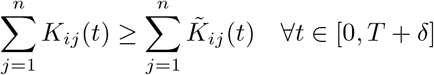

and, by identical arguments (using 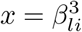 and 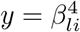 in Lemma A.8)

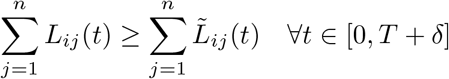

as required.

### A.10 Lemma A.10

#### Lemma A.10.

*Suppose that*

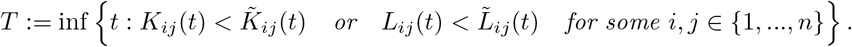

*Then, for any δ >* 0, *there exists some t* ∈ (*T, T* + *δ*) *and some real parameters* 0 ≤ *κ* ≤ *η* ≤ 1 *such that*

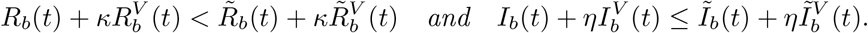

**Proof:** Firstly, note that by the definition of *T*, for each *δ >* 0, there must exist *i, j* ∈ *{*1, …, *n}* and *t* ∈ (0, *δ*) such that

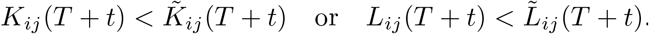

That is, there is some *b* ∈ *{*1, .., *n}* such that

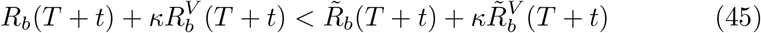

where

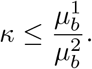

Note that

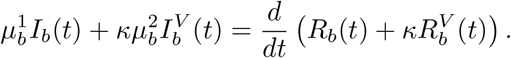

Now, define

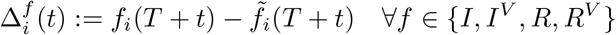

and

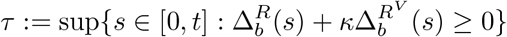

which exists as 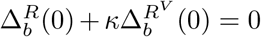. Note that *τ < t* by (45). Note also that by continuity, it is necessary that

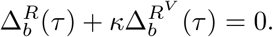

Now, by the mean value theorem (as 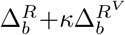 is continuously differentiable), there exists an *s* in the non empty interval (*τ, t*) such that

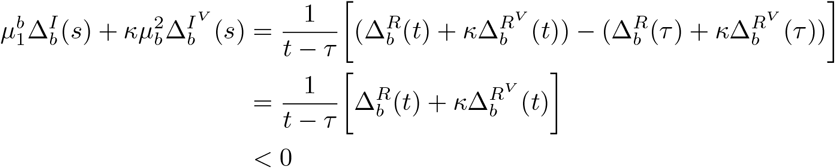

while also

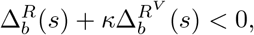

by definition of *τ*. Thus, defining 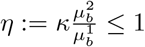,

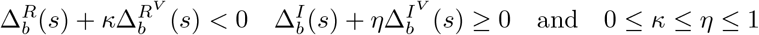

as required.

### A.11 Lemma A.11

#### Lemma A.11.

*Consider two non-negative functions A*(*t*) *and B*(*t*) *such that B*(*t*) *is non-decreasing and differentiable with a Lebesgue integrable derivative B*′(*t*) *satisfying*

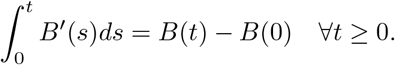

*Suppose further that for each T* ≥ 0, *one can partition the interval* [0, *T*] *into a finite number of subintervals* 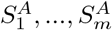 *and* 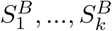 *such that*

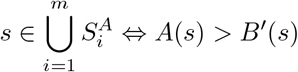

*Then, there exists a unique function χ*(*t*) *for t* ≥ 0 *such that*

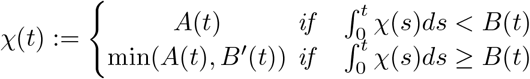

**Proof:** *χ* can be constructed for each of the subintervals 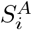 and 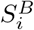. Note first that,

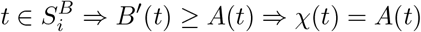

Now, suppose that 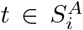 for some *i*. Then, as 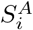 is an interval, one can suppose 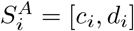. Define

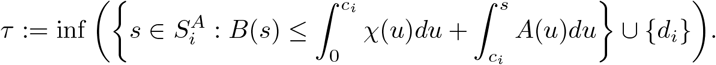

If *τ* = *d*_*i*_, then one has (uniquely) *χ*(*t*) = *A*(*t*) in 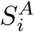. Otherwise, one has (again uniquely)

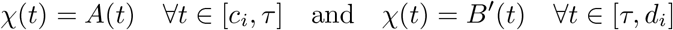

Uniqueness can be demonstrated as follows. If *χ*(*t*) = *B*′(*t*) for some *t* ∈ [*c*_*i*_, *τ*], then it is necessary (as *A*(*t*) *> B*′(*t*) so *χ*(*t*) ≠ *A*(*t*) in this case)

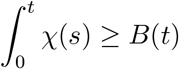

As *A*(*t*) ≥ *B*′(*t*) in 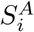, so *χ*(*t*) is bounded by *A*(*t*), the previous inequality can be extended to give

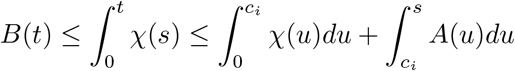

which contradicts the definition of *τ*. A similar argument stands to prove uniqueness in [*τ, d*_*i*_].

Thus, *χ* is uniquely defined in each of the finite number of intervals and hence in [0, *T*] for each *T* and hence, it is uniquely defined for all *t* as required.

## B Results on the SIR Equations

This section presents a variety of results on the SIR equations which are used in the proofs of the theorems in this paper. Many of them are well-known and widely used in the literature, but this appendix aims to provide a source of formal definitions and proofs of these results.

Before the results can be proved, it is necessary to establish two lemmas on differential equations.

### B.1 Lemma B.1

#### Lemma B.1.

*Suppose that H*(*t*) *is a continuous non-negative n* × *n matrix for t* ≥ 0 *and that* ***a*** ∈ ℜ ^*n*^. *Then, suppose that a function* ***u*** : ℜ → ℜ ^*n*^ *satisfies*

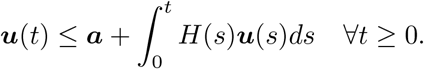

*Then*,

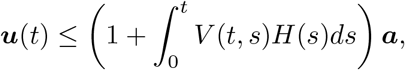

*where the matrix V* (*t, s*) *satisfies*

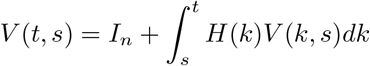

*and I*_*n*_ *is the n* × *n identity matrix*.

**Proof:** This theorem is a special case of the theorem proved in Chandra and Davis (1976) where (in the notation of Chandra and Davis (1976)), *x, y* and *z* have been replaced by *t, s* and *k* respectively, *G*(*t*) has been set to be the identity matrix and *x*^0^ has been set to zero.

### B.2 Lemma B.2

#### Lemma B.2.

*Consider a continuous, time-dependent, matrix A*(*t*) *which satisfies*

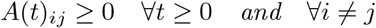

*and a constant matrix B that satisfies*

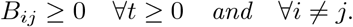

*Then, suppose that each element of A*(*t*) *is non-increasing with t and that*

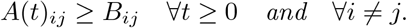

*Moreover, define a non-negative initial condition* ***v*** *and suppose that* ***y*** *and* ***z*** *solve the systems*

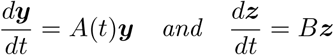

*with*

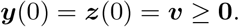

*Then*,

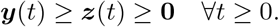

**Proof:** To begin, define

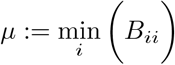

so that, defining

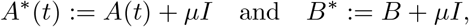

where *I* is the identity matrix, *A*^*^ and *B*^*^ are non-negative matrices. Moreover, note that

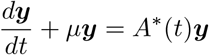

and so

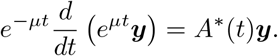

Thus, define

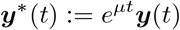

so

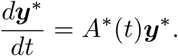

Similarly, defining

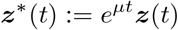

gives

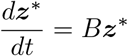

while, moreover,

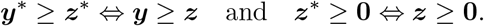

Thus, it is simply necessary to prove that the results of this lemma hold when *A*(*t*) and *B* are non-negative matrices.

Now, it is helpful to note that, as the off-diagonal entries of *A*(*t*) and *B* are non-negative, the two differential systems are totally positive Schwarz (1970). Thus, in particular, as ***v*** is non-negative,

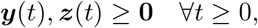

which proves one of the required inequalities. Now, one can also note that

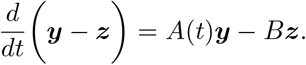

As *A*(*t*) is assumed to be non-negative, and ***y*** is non-negative,

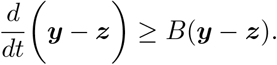

Defining ***ζ*** := ***z*** − ***y*** and integrating gives

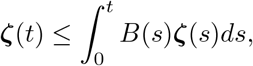

noting that ***ζ*** = **0**. Hence, by Lemma B.1, one has

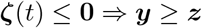

as required.

### B.3 Lemma B.3

#### Lemma B.3.

*Define the set of functions*

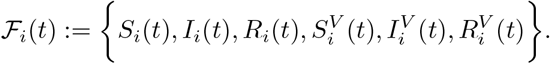

*Then, for all t* ≥ 0 *and i* ∈ *{*1, …, *n}*,

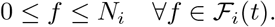

**Proof:** Noting that

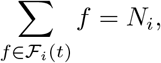

it is simply necessary to show that (for each *t* and *i*)

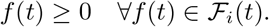

Now, note that

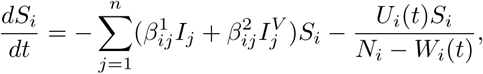

which means

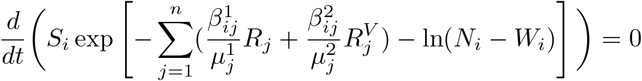

and hence (using the initial conditions)

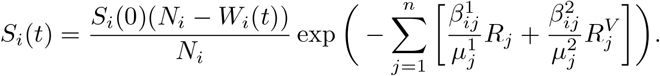

As *W*_*i*_(*t*) ≤ *N*_*i*_ by construction, this means that

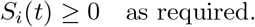

Now, note that

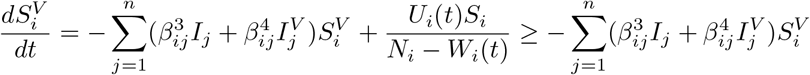

so that

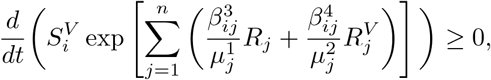

which means (as 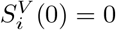)

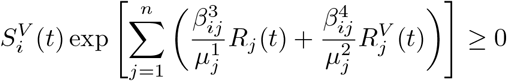

and hence

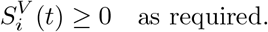

Now, define the vector

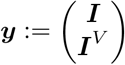

Then, one can rewrite the equations for *I*_*i*_ and 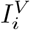 in the form

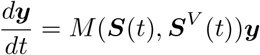

for some matrix *M*, where, from the previous results

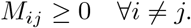

Thus, from Lemma B.2,

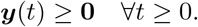

Then,

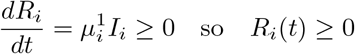

and similarly,

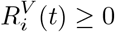

and so the proof is complete.

### B.4 Lemma B.4

#### Lemma B.4.

*For each i*,

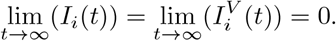

**Proof:** Firstly, suppose

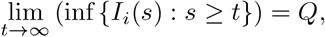

noting this infimum exists as *I*_*i*_ is bounded below by 0, and the limit exists as the sequence of infima given *s* ≤ *t* is non-decreasing and bounded above by *N*_*i*_. If *Q* ≠ 0, there exists some *m >* 0 and some *t* such that for all *s* ≥ *t*

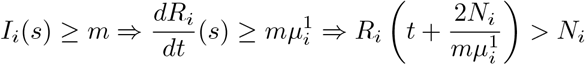

which contradicts Lemma B.3. Thus, *Q* = 0 and so there exists some sequence *t*_*n*_ such that

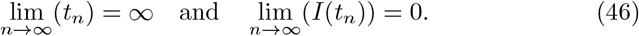

Now note that *S*_*i*_(*t*) is non-increasing and bounded and that *R*_*i*_(*t*) and 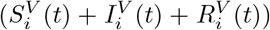 are non-decreasing and bounded. Thus, their limits as *t* → ∞ must exist and be finite, so in particular

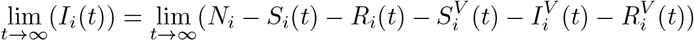

must exist. Thus, by (46), the only possible limit is 0 so

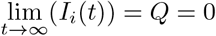

as required. By noting that 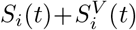 is non-increasing and that *I*_*i*_(*t*)+*R*_*i*_(*t*) and 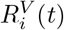 are non-decreasing, an identical argument shows that

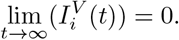

### B.5 Lemma B.5

#### Lemma B.5.

*Suppose that I*_*i*_(*t*) *>* 0 *for some t* ≥ 0 *and some i* ∈ {1, …, *n* }. *Then*,

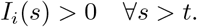

*An analogous result holds for* 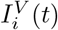.

**Proof:** Note that

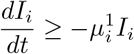

and so

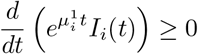

which means, for any *s > t*

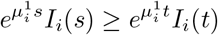

and hence

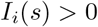

as required. The same argument then works for 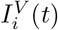 as well (with a 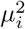 instead of a 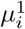).

### B.6 Lemma B.6

#### Lemma B.6.

*Define*

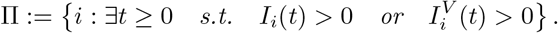

*Moreover, define*

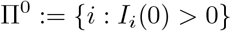

*and the n by n matrix M by*

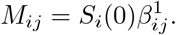

*Then, define the connected component C of* Π^0^ *in M as follows. The index i* ∈ {1, …, *n* } *belongs to C if any only if there is some sequence a*_1_, …, *a*_*k*_ *such that*

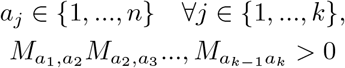

*and*

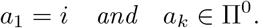

*Then*,

a. *i* ∈ *C* ⟹ *I*_*i*_(*t*) *>* 0 ∀*t >* 0.
b. Π = *C* ∪ Π^0^.

*Thus, in particular*,

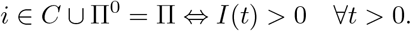

**Proof: (a):** The proof will proceed by induction. For *k* ≥ 1, define *P*^*k*^ is the set of elements of *C* that are connected to an element of Π^0^ by a sequence of length at most *k*. Then, note that

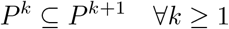

and

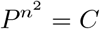

as there are *n*^2^ elements in *M*. (Thus, if *i ∈ C* then there must be a sequence of length at most *n*^2^ connecting *i* with an element in Π^0^ as any loops can be ignored.)

The inductive hypothesis is that

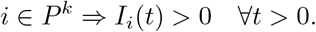

The explanation of the base case will be left until the end of the proof. Suppose that this claim holds for some *k* ≥ 0. If *P*^*k*+1^ = *P*^*k*^, then

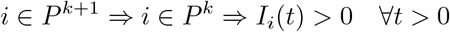

and so the inductive step is complete. Otherwise, consider any *i* ∈ *P*^*k*+1^*/P*^*k*^. Then, there exists some *j* such that

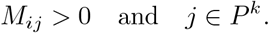

Thus, by continuity, for sufficiently small *τ*,

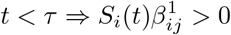

and indeed, by Boundedness Theorem, there exists some *χ >* 0 such that

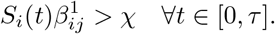

Now, choose any *ϵ* ∈ [0, *τ*]. By Boundedness Theorem, *I*_*i*_(*t*) achieves is bounded and achieves its maximum, *θ*_*ϵ*_ in the interval [0, *ϵ*]. Moreover, *θ*_*ϵ*_ *>* 0 as *I*_*i*_(*t*) *>* 0 in (0, *ϵ*) by assumption. Thus, by continuity, there exists some non-empty region (*δ*_*ϵ*_, Δ_*ϵ*_) such that

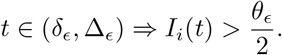

Thus, in particular

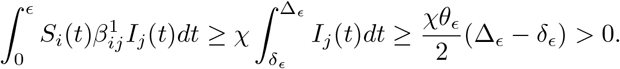

Now, note that

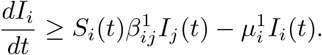

Suppose for a contradiction that *I*_*i*_(*t*) = 0 for all *t* ∈ [0, *ϵ*]. Then,

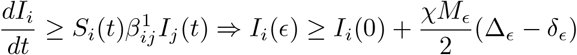

and hence,

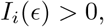

which is a contradiction. Thus, there exists a *t* ∈ [0, *ϵ*] such that *I*_*i*_(*t*) *>* 0 and hence, by Lemma B.5,

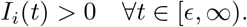

Thus, as *ϵ* was any constant in the region (0, *τ*), and *τ >* 0, this means that

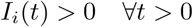

as required.

Finally, note that the base case *k* = 1 can be proved in exactly the same way, except now *j* ∈ Π^0^ (but this still means that *I*_*j*_(*t*) *>* 0 for all *t >* 0 by Lemma B.5), and so **(a)** has been proved.

**(b)**: The previous work has shown that

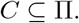

Hence, as clearly Π^0^ ⊆ Π, this means that

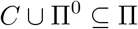

and so it suffices to prove that

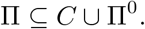

That is, it suffices to prove

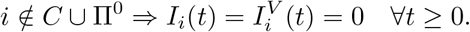

To check that this solution satisfies the equations, one notes that, in this case, if *i* ∉ *C* ∪ Π^0^, then

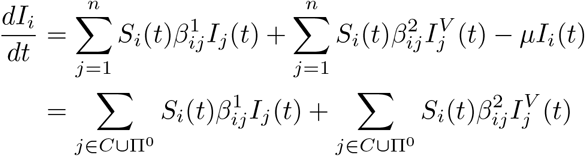

and, similarly,

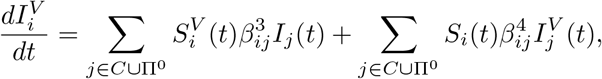

as 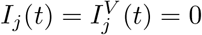 for all *j* ∉ *C* ∪ Π^0^.

Now, suppose that *i* ∉ *C* Π^0^ and *j* ∈ *C* ∪ Π^0^. Then, by definition of *C*, this means that

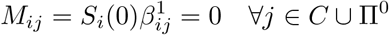

and hence, as *S*_*i*_ is non-increasing and non-negative

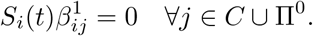

Now, as 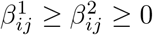, this means that

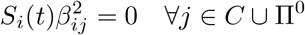

so that

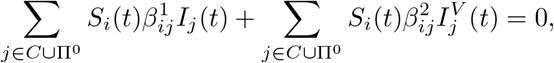

which means

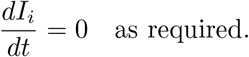

Moreover, as 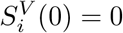, it is necessary that

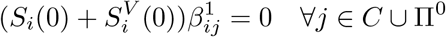

so, as 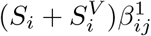 is non-increasing and non-negative

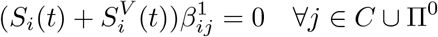

and hence, as *S*_*i*_(*t*) is non-negative

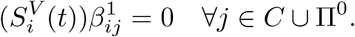

Thus, as 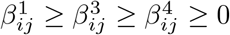, one has

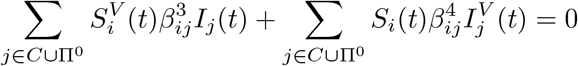

and hence

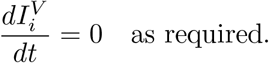

Then, one can separately solve the system for all *j* ∈ *C* ∪ Π^0^ as the equations will now be independent of any indices *i* ∉ *C* ∪ Π^0^ (as they only depend on these indices via the *I*_*i*_ and 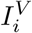 terms, which are identically zero). Thus, by the uniqueness of solution, one must have

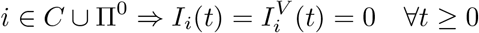

and hence part **(b)** is proved. Thus, the lemma has been proved.

### B.7 Lemma B.7

#### Lemma B.7.

*Consider a set C* = [*a*_1_, *b*_1_] × [*a*_2_, *b*_2_]× … × [*a*_*n*_, *b*_*n*_] *that is a Cartesian product of real intervals. Suppose that f* : ℜ^*n*^→ ℜ *is differentiable with bounded derivatives in C. Then, f is Lipschitz continuous on C - that is, there exists some L >* 0 *such that*

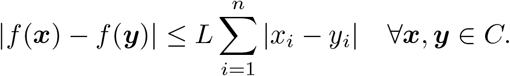

**Proof:** Note that, by assumption, for each *i*,

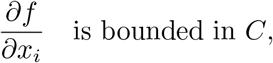

so define the global bound for all *i* to be M. Choose some ***x, y*** ∈ *C*. Define the points ***p***^*k*^ ∈ *C* for *k* = 0, 1,, *n* by

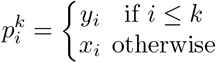

and define the curve *γ*_*i*_ to be the straight line joining the point ***p***^*i*−1^ to the point ***p***^*i*^. As *C* is a product of intervals, the *γ*_*i*_ lie entirely in *C*.

Define Γ to be the union of the curves *γ*_*i*_, so that Γ joins ***p***^0^ = ***x*** to ***p***^*n*^ = ***y***. Then

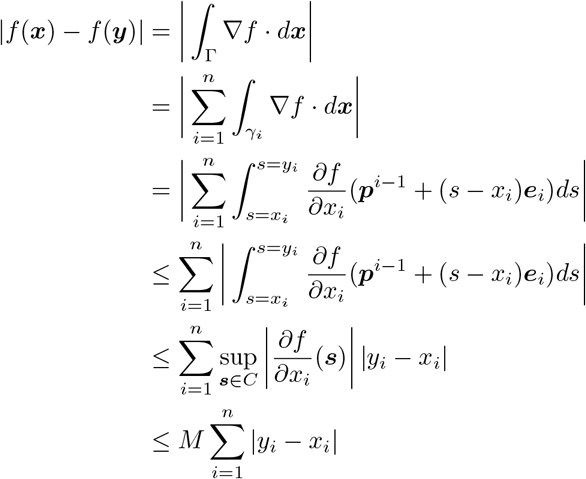

where ***e***_*i*_ is the ith canonical basis vector. Hence, the required Lipschitz continuity holds with *M* = *L*.

### B.8 Lemma B.8

#### Lemma B.8.

*Define the set of functions*

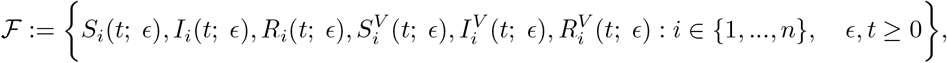

*where for each fixed ϵ, these functions solve the model equations with parameters*

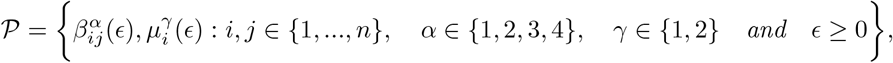

*initial conditions*

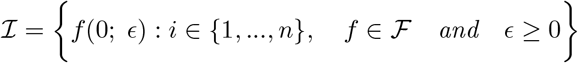

*and vaccination policy* ***U*** (*t*; *ϵ*). *Suppose that*

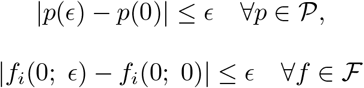

*and that*

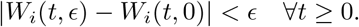

*Moreover, suppose that for each i* ∈ *{*1, …, *n} and ϵ* ≥ 0,

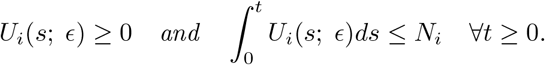

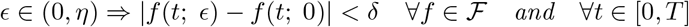

**Proof:** To begin, it is helpful to note that, by Lemma B.3,

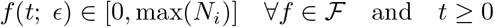

and that, by assumption on the feasibility of *U*_*i*_

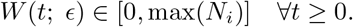

Moreover, as the parameter values converge, it can be assumed that

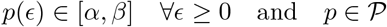

for some *α, β* ≥ 0. Moreover, it can be assumed that, as each 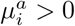, there is some *γ >* 0 such that 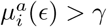 for all *ϵ* ≥ 0.

However, there is no condition on the maximal difference (at a point) between *U*_*i*_(*t*; *ϵ*) and *U*_*i*_(*t*; 0). To avoid this problem, it is helpful to consider the variable 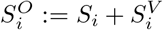 instead of 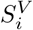. Then, the equations for *S*_*i*_ and 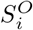 can be written as

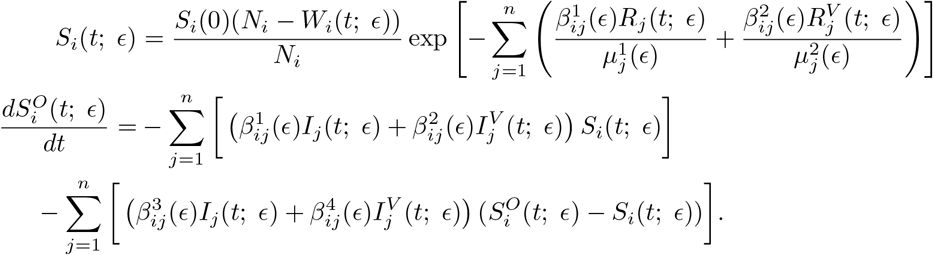

Then, one can define

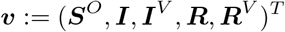

and ***p***(*ϵ*) to be a vector of the elements of 𝒫 at some *ϵ* ≥ 0. Then, (substituting for ***S***), the model equations can be written in the form

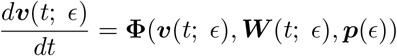

where Φ is a smooth function. Thus, from Lemma B.7, there exists some constant *L* such that, for ***v, W*** and ***p*** within the closed bounded feasible set of values and any *j* ∈ *{*1, …, 5*n}*,

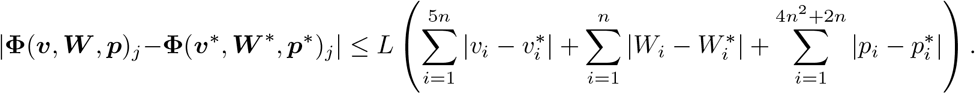

Thus, in particular, this means that

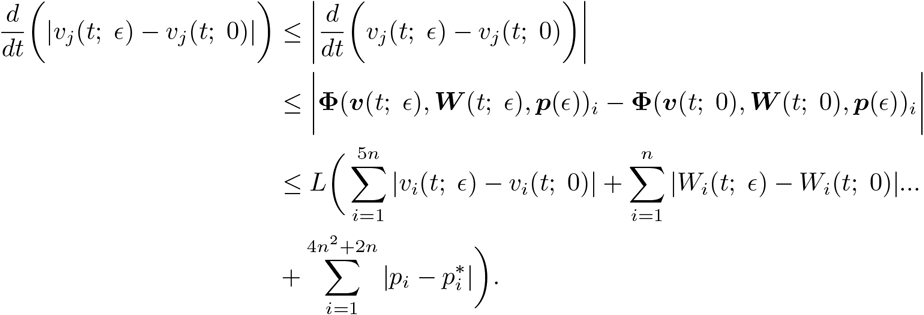

Now, adding these 5*n* inequalities together, one seems that

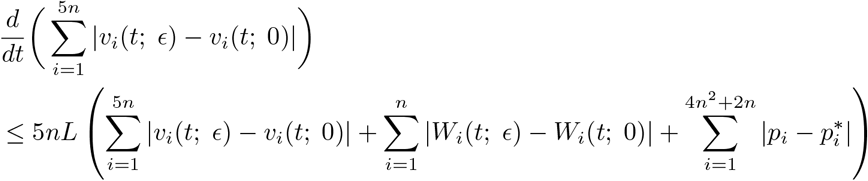

and hence

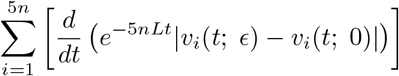

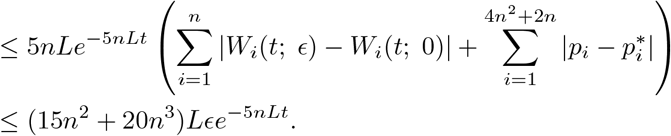

Thus, integrating (and using the fact that the initial conditions differ by at most *ϵ*)

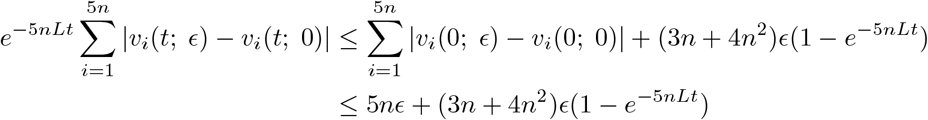

which means

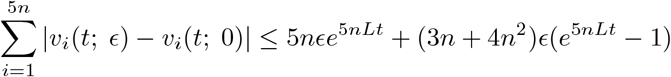

and hence, for each *i* ∈ *{*1, …, 5*n}*

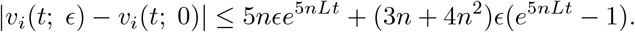

The right-hand side is non-decreasing in *t* (as *L >* 0) so, taking

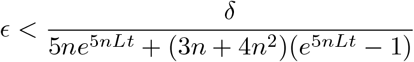

ensures that the required inequalities hold for ***I, I***^*V*^, ***R*** and ***R***^*V*^ for all *s* ≤ *t*. Now, note also that *S*_*i*_(*t*; *ϵ*) is a smooth function of *W*_*i*_(*t*; *ϵ*), ***v***(*ϵ*), *S*_*i*_(0; *ϵ*) and ***p*** so that there exists an *L*′ such that

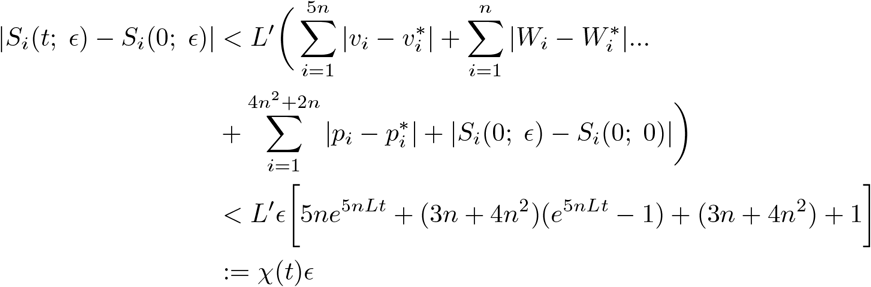

and so, as *χ*(*t*) is non-decreasing in *t*, taking

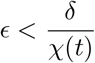

gives the required inequalities for ***S*** for all times *s* ≤ *t*. Finally, note that

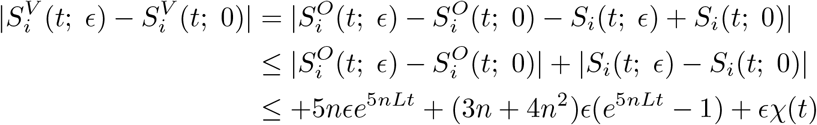

and so, as the right-hand side is increasing in *t*, taking

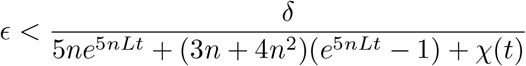

gives the required inequalities for ***S***^*V*^ for all times *s ≤ t* and hence completes the proof.

